# Olfactory Function in Preclinical Alzheimer’s Disease: A Multi-Cohort Study

**DOI:** 10.64898/2026.11.03.26367202

**Authors:** Benoît Jobin, Yasmine Salman, Kamar Bouchoucha, Lara Huyghe, Lisa Quenon, Lise Colmant, Thomas Gérard, Vincent Malotaux, Laurence Dricot, Paul Dulieu, Adrian Ivanoiu, Renaud Lhommel, Anabelle Decottignies, Bernard Hanseeuw, Caroline Huart, The PREVENT-AD Research Group

**Author notes:** Corresponding author: Benoît Jobin, 114 Sixteenth St., Charlestown, MA 02129. Co-first authorship (Benoît Jobin, Yasmine Salman). Co-last authorship (Bernard Hanseeuw, Caroline Huart).

## Abstract

**Introduction:** Olfactory impairment is an early feature of Alzheimer’s disease (AD). However, it is unclear if olfactory performance reflects early pathological changes during preclinical AD and/or predicts subsequent clinical progression among characterized cognitively normal (CN) individuals. We aimed to determine whether odor identification differs by amyloid status, relates to medial temporal lobe (MTL) tau burden and morphometry, and predicts longitudinal conversion to mild cognitive impairment (MCI).

**Methods:** We analyzed data from 257 cognitively normal (CN) older adults enrolled in PREVENT-AD (n = 217) and UCLouvain (n = 40), including 76 amyloid-positive (CN A+) and 181 amyloid-negative participants (CN A–). Participants completed olfactory testing, underwent tau-PET imaging, and structural MRI.

**Results:** CN A+ participants had significantly lower odor identification performance than CN A− participants, with no evidence of cohort heterogeneity. Tau-PET associations with odor identification differed by cohort. After whole-brain FDR correction and exclusion of extreme values, parahippocampal tau burden was negatively associated with odor identification score in UCLouvain ([18F]MK-6240), an association that was not found in PREVENT-AD ([^18^F]flortaucipir). No morphometric association survived multiple comparison correction. In 250 participants with longitudinal follow-up, each 10-percentage-point higher odor identification score was associated with a 16% lower hazard of incident MCI, independent of baseline global cognition and APOE ε4-carrier status.

**Discussion:** Odor identification was lower in preclinical AD and associated with incident MCI independently of baseline global cognition and APOE ε4-carrier status. Its association with parahippocampal tau burden differed between cohorts, supporting further investigation using harmonized imaging methods.

**Clinical trial registration:** Not applicable; this was an observational cohort study.

## 1. Background

Alzheimer’s disease (AD) is a progressive neurodegenerative disease characterized by amyloid beta (Aβ) plaques and hyperphosphorylated tau protein aggregates, leading to neurodegeneration and gradual cognitive decline that ultimately culminates in dementia ^1,2^. The hypothesis suggests that Aβ precedes and exacerbates tau accumulation, leading to neurodegeneration ^3,4^. Preclinical AD can be detected in vivo decades before the dementia stage in cognitively normal (CN) individuals using positron emission tomography (PET), spinal tap extraction of cerebrospinal fluid (CSF), or more recently developed blood-based assays ^5–7^.

The recent approval of anti-amyloid monoclonal antibodies for symptomatic early AD ^8–10^ has underscored the growing need to develop preventive strategies targeting earlier, preclinical stages of the disease. Yet, identifying CN individuals at increased risk remains challenging at the population level. Although blood-based biomarkers offer a less invasive alternative to CSF and PET, recent Alzheimer’s Association guidelines emphasize that their clinical use should currently be reserved for individuals with objective cognitive impairment, such as Mild cognitive impairment (MCI), rather than for CN individuals.^11^ This gap highlights the need for complementary, cost-effective, and scalable presymptomatic screening tools to identify CN individuals at highest risk of cognitive decline, particularly those with combined amyloid and tau pathology.

Olfactory testing could offer an accessible and biologically grounded approach to preclinical AD screening. The neural circuits of the olfactory system are among the first structures damaged by tau pathology at Braak stage I (i.e., the olfactory bulb and entorhinal cortex) ^12,13^ and cerebral atrophy (i.e., amygdala and hippocampus) ^14–17^. The capacity to identify odors declines early in the course of AD, being already impaired before objective memory deficits in individuals reporting subjective cognitive decline (SCD) ^18,19^. Lower olfactory performance has been associated with lower episodic memory ^20,21^, longitudinal cognitive decline ^22^, smaller medial temporal lobe (MTL) volumes ^23–25^ and tau pathology burden measured by PET imaging, particularly the hippocampus and entorhinal cortex ^26,27^.

One study reported that poorer odor identification was associated with greater tau burden in the medial temporal lobe and hippocampus among CN individuals who tested positive for amyloid pathology (CN A+) and patients with MCI ^28^. A recent study from our group, conducted in an age-matched cohort, replicated this finding by showing impaired odor identification, but preserved odor discrimination and detection threshold, in CN individuals^29^. In the current study, we aimed to examine whether olfactory performance reflects early pathological changes in the MTL during the preclinical AD phase in CN A+ individuals. More specifically, we aimed to (1) compare olfactory function between CN individuals with and without (CN A–) amyloid pathology; (2) assess whether olfactory scores are associated with tau burden and the morphometry of MTL subregions; and (3) assess the predictive value of odor identification on longitudinal conversion to MCI. We analyzed data from the PREVENT-AD and UCLouvain cohorts, which combine neuropsychological testing, standardized olfactory measures, structural MRI, and tau PET imaging. This multimodal design enabled us to evaluate olfactory performance as a potential preclinical marker of AD.

## 2. Methods

### 2.1 Participants

#### 2.1.1 PREVENT-AD Cohort

The PREVENT-AD cohort is a longitudinal observational study based at the Douglas Mental Health University Institute in Montréal, Canada. Since 2011, it has enrolled cognitively unimpaired older adults at increased risk of AD because of a parental or multiple-sibling history of AD dementia. Eligible participants were ≥60 years old or 55–59 years old if ≤15 years of the youngest age at onset in their affected first-degree relative. Participants had at least six years of education, were fluent in French and/or English, had a study partner, and underwent annual cognitive follow-up. At enrollment, participants completed a health review, neurological and physical examination, blood sampling, structural MRI, and cognitive assessment including the Montreal Cognitive Assessment (MoCA) ^30^ , and Clinical Dementia Rating (CDR) ^31^. Participants were excluded if clinical consensus determined probable MCI, major neurological or psychiatric illness, substance abuse, or contraindications to MRI or lumbar puncture. Full PREVENT-AD procedures have been described previously ^32,33^. PREVENT-AD did not apply a specific rhinological exclusion criterion.

We analyzed data collected from participants who underwent olfactory, MRI, and tau-PET examinations between October 2013 and October 2023. For each participant, the baseline analytic visit was defined as the olfactory assessment–PET imaging pair with the smallest absolute time interval. Amyloid status was determined using [^18^F]NAV4694 PET and quantified on the Centiloid scale; participants were considered amyloid-positive when the Centiloid value exceeded 20 ^34^. Regional tau burden was assessed using [^18^F]flortaucipir PET (FTP-PET), and structural MRI was used to derive medial temporal lobe regions of interest (ROI) morphometric measures. Cognitive and clinical outcomes were assessed longitudinally through annual follow-up visits. From the PREVENT-AD cohort, 159 CN A– and 58 CN A+ participants were included. Data used in preparation of this article were obtained from the PREVENT-AD Registered repository available at https://registeredpreventad.loris.ca. The Investigators of the PREVENT-AD program contributed to the design and implementation of PREVENT-AD and/or provided data but did not participate in data analysis or writing of this report.

#### 2.1.2 UCLouvain Cohort

Older adult participants were enrolled in a study conducted at UCLouvain, Brussels, Belgium. Patients were recruited through the memory clinic of the Cliniques Universitaires Saint-Luc, whereas volunteers were recruited from ongoing research studies and through mailbox announcements distributed in the hospital’s neighborhood. The volunteer sample was enriched for APOE ε4 carriers, who represented approximately half of the volunteers included in the study. Thirty-eight participants were drawn from the UCL-2018-0034/73-94 study, with data collected between March 2019 and January 2025, and the remaining participants were drawn from the UCL-2011-001756-12 study, with data collected between 2012 and 2021. Exclusion criteria for both patients and volunteers included focal brain lesions, epileptic seizures, major depression or psychiatric conditions, alcohol or drug abuse, chronic rhinosinusitis, and head trauma.

Participants underwent a comprehensive neuropsychological assessment, amyloid PET imaging or spinal tap, and structural MRI. A neuropsychological follow-up assessment was proposed every two years. A subset of participants also underwent tau-PET imaging with [^18^F]MK-6240 (13 CN A− ; 17 CN A+ participants). The comprehensive neuropsychological assessment covered four cognitive domains: memory, language, executive functions, and visuospatial abilities. For each cognitive domain, three measures were obtained and converted into z-scores. A cognitive domain was considered impaired when the corresponding z-score fell below −1.5 SD relative to the mean performance of an independent reference group of 32 individuals who remained cognitively stable over an eight-year follow-up (for further details, see ^35^). Participants were excluded if they were classified as having MCI, with at least one cognitive domain impaired.

We analyzed data collected from participants who underwent olfactory, MRI, and PET assessments. Amyloid status was determined from amyloid-PET or cerebrospinal-fluid biomarkers. Regional tau-PET SUVR analyses included 13 CN A− and 17 CN A+ participants. Visual tau reads (examined by RL and TG) were available for 13 CN A− and 18 CN A+ participants; tau positivity was defined as a Braak-like stage greater than 0. Three UCLouvain participants with an observed cognitive-domain z-score below −1.5 before olfactory testing were excluded from all analyses. In total, 40 CN participants were included in the primary cross-sectional group comparisons and 34 had longitudinal follow-up available for the incident-MCI analyses. UCLouvain de-identified data are available upon request to the authors. In total, 257 participants were included, comprising 181 CN A– and 76 CN A+.

### 2.2 Olfactory assessment

#### 2.2.3 PREVENT-AD Cohort

The University of Pennsylvania Smell Identification Test (UPSIT) ^36^ was used to measure odor identification in the PREVENT-AD cohort. The UPSIT is a 40-item scratch-and-sniff test where the participant has to identify a target odor among a set of four written choice options.

#### 2.2.2 UCLouvain Cohort

In the UCLouvain cohort, the Sniffin’ Sticks test battery was used to assess olfactory detection threshold, discrimination of odors, and odor identification ^37^. Both the UPSIT and the Sniffin’ Sticks battery have been shown to be strongly correlated ^38^ and both showed good psychometric performance ^36,39^.

To harmonize odor-identification performance across cohorts, UPSIT scores were divided by the maximum possible score of 40, whereas Sniffin’ Sticks identification scores were divided by the maximum possible score of 16. For the exploratory UCLouvain analyses, discrimination and detection-threshold scores were similarly divided by their maximum possible score of 16. The resulting proportional scores were expressed as percentages of the maximum possible score, with higher scores indicating better olfactory performance.

### 2.3 Neuroimaging Acquisition

The complete neuroimaging acquisition methodology is described in the Supplementary Materials.

#### 2.3.1 PREVENT-AD Cohort

MRI scans were acquired at the Cerebral Imaging Center of the Douglas Mental Health University Institute in Montréal. During Phase 1 (2011–2017), T1-weighted neuroimaging was performed using a 3T Siemens Tim Trio scanner. In Phase 2 (2019 onward), MRI acquisition resumed following an upgrade to a 3T Siemens Prisma Fit scanner.

Tau-PET was performed at the McConnell Brain Imaging Centre of the Montreal Neurological Institute using a brain-dedicated Siemens/CTI high-resolution research tomograph (HRRT), with a spatial resolution of approximately 2.4 mm at the center of the field of view. Participants received approximately 370 MBq of the first-generation tau tracer [^18^F]Flortaucipir, and four 5-minute frames were acquired 80–100 minutes post-injection. Frames were realigned, averaged, and registered to each participant’s T1-weighted MRI. Standardized uptake value ratio images were then generated using the inferior cerebellar gray matter as the reference region.

#### 2.3.2 UCLouvain Cohort

MRI scans were acquired at the Cliniques Universitaires Saint-Luc in Brussels, Belgium. Before October 2019, 3D T1-weighted images were acquired on a 3T MRI scanner (Achieva, Philips Healthcare) using a 32-channel phased-array head coil. Following the replacement of this scanner, acquisitions were performed on a 3T Signa™ Premier system (GE Healthcare) equipped with a 48-channel phased-array head coil.

Ninety minutes after intravenous administration of [^18^F]MK-6240 (target activity 185 ± 5 MBq) a 30-minute dynamic list-mode acquisition was performed on a Philips Vereos digital PET-CT (Philips Healthcare). PSF correction and 1mm re-slicing was applied using manufacturer’s algorithm. The cerebellar gray matter was used as a reference to generate [^18^F]MK-6240 SUVR.

### 2.4 Neuroimaging processing

In both cohorts, structural MRI data were processed using FreeSurfer, including subcortical segmentation and cortical parcellation according to the Desikan–Killiany atlas ^40^. FreeSurfer v7.2 was used for the UCLouvain cohort and v8.0 for PREVENT-AD. Compared with v7.2, v8.0 incorporates deep learning-based skull stripping and segmentation, together with refinements to cortical surface reconstruction, particularly in the entorhinal cortex.

Tau-PET preprocessing followed similar cohort-specific pipelines: In PREVENT-AD, images were processed using a standard Villeneuve’s Lab in-house pipeline (https://github.com/villeneuvelab/vlpp). In UCLouvain, tau-PET data were processed using the PetSurfer pipeline, a set of tools within FreeSurfer for integrated MRI-PET analysis ^41^. In both cohorts, tau-PET frames were realigned, averaged, and co-registered to the corresponding T1-weighted MRI. In PREVENT-AD, inferior cerebellar gray matter was used as the reference region to generate [^18^F]Flortaucipir SUVR, whereas in UCLouvain, whole cerebellar gray matter was used as the reference region to generate [^18^F]MK-6240 SUVR.

We extracted SUVR and morphometric measures for prespecified ROIs, which included the amygdala and hippocampus (grey matter volume, GMV) and the entorhinal cortex and parahippocampal gyrus (cortical thickness). These MTL ROIs are known to be part of the primary and secondary olfactory areas ^42–44^. Bilateral ROI values were computed by averaging left and right SUVR and cortical thickness values, and by summing left and right GMV. Whole-brain analyses were exploratory.

### 2.5 Statistical Analyses

To compare olfactory performance across CN A− and CN A+ groups, we used ANCOVA models adjusted for cohort, age, sex, education, and APOE ε4-carrier status. Because education was measured in years in PREVENT-AD and as an ordinal category in UCLouvain, education was centered within each cohort and entered using separate cohort-specific covariates. An amyloid-status-by-cohort interaction tested whether the adjusted group difference varied between cohorts. All tests were two-sided, with *p* < .05 considered statistically significant. In exploratory UCLouvain analyses, discrimination and threshold scores were analyzed using separate ANCOVA models with the same covariates.

Regarding neuroimaging associations, we first fit combined-cohort linear models relating normalized odor identification scores to regional tau-PET or morphometric measures standardized within cohort. These models estimated a common biomarker association across cohorts and were adjusted for cohort, amyloid status, age, sex, education, and APOE ε4-carrier status. We then added a regional biomarker-by-cohort interaction to test whether the association differed between PREVENT-AD and UCLouvain. When an interaction survived FDR correction, cohort-specific slopes from the interaction model were examined as follow-up estimates. Subcortical-volume models were additionally adjusted for eTIV. Tau-PET models additionally included global cortical tau and its interactions with cohort and amyloid status. Coefficients represent percentage-point differences in odor-identification score per 1 within-cohort SD higher biomarker value. Regional biomarker-by-amyloid and regional biomarker-by-cohort-by-amyloid interactions were exploratory. FDR correction was applied across the four prespecified MTL ROIs and, separately, across 42 atlas-wide regions for each modality and statistical estimand. Extreme-value sensitivity analyses excluded observations with regional neuroimaging biomarker values beyond ±3 within-cohort SDs. In UCLouvain, an exploratory adjusted model examined odor identification according to visual tau positivity, defined as a Braak-like stage greater than 0.

Incident MCI was examined in both cohorts together using Cox models with separate baseline hazard functions for PREVENT-AD and UCLouvain. Models were adjusted for amyloid status, age, sex, education, APOE ε4-carrier status, and baseline global cognition measured using the MoCA in PREVENT-AD and the MMSE in UCLouvain. Hazard ratios were estimated per 10-percentage-point higher odor identification score. Odor identification-by-amyloid-status and odor identification-by-cohort interactions were tested using likelihood-ratio tests. Proportional hazards assumptions were assessed using Schoenfeld residuals. Kaplan–Meier curves based on cohort-specific odor-identification tertiles were used for descriptive visualization. All tests were two-sided, with *p* < .05 considered statistically significant unless FDR correction was specified.

## 3. Results

Participant characteristics are presented in Table 1. In PREVENT-AD, CN A+ participants had lower unadjusted UPSIT scores than CN A− participants (*p* = .027), whereas demographic variables did not differ by amyloid status. In UCLouvain, CN A+ participants were older (*p* = .025) and had lower MMSE scores (*p* = .022) than CN A− participants; Sniffin’ Sticks scores did not differ significantly by amyloid status.

**Table 1.** Participant Characteristics and Medial Temporal Lobe Biomarkers.

|  | **PREVENT-AD<sup>a</sup>** |  |  | **UCLouvain<sup>b</sup>** |  |  |
|---|---|---|---|---|---|---|
| **Mean (SD)** | **CN A−** | **CN A+** | ***p*** | **CN A−** | **CN A+** | ***p*** |
| ***Demographics*** |  |  |  |  |  |  |
| n | 159 | 58 |  | 22 | 18 |  |
| Age (y) | 65.43 (5.07) | 66.91 (5.46) | .074 | 69.90 (6.44) | 74.49 (6.00) | **.025** |
| Sex (%F) | 71.7 | 67.2 | .614 | 36.4 | 55.6 | .339 |
| Education | 15.64 (3.44) | 14.91 (3.02) | .133 | 2.95 (0.21) | 2.89 (0.47) | .590 |
| APOE ε4 carrier (%) | 30.2 | 69.0 | <.001 | 36.4 | 61.1 | .203 |
| ***Cognition*** |  |  |  |  |  |  |
| MoCA | 28.16 (1.52) | 28.21 (1.50) | .833 |  |  |  |
| MMSE |  |  |  | 29.36 (0.58) | 28.78 (0.88) | **.022** |
| ***Olfaction*** |  |  |  |  |  |  |
| UPSIT identification | 34.82 (4.68) | 32.72 (6.49) | **.027** |  |  |  |
| Sniffin' Sticks identification |  |  |  | 12.00 (2.45) | 10.17 (3.54) | .072 |
| Sniffin' Sticks discrimination |  |  |  | 10.41 (2.99) | 10.33 (3.01) | .937 |
| Sniffin' Sticks threshold |  |  |  | 6.56 (3.12) | 5.51 (3.12) | .296 |
| ***Morphometry*** |  |  |  |  |  |  |
| Amygdala volume (mm³) | 3368.69 (359.77) | 3304.91 (415.12) | .315 | 3160.65 (450.35) | 3005.90 (471.58) | .299 |
| Hippocampal volume (mm³) | 7947.39 (812.27) | 7705.58 (977.59) | .104 | 6740.51 (923.49) | 5843.14 (704.71) | **.001** |
| Entorhinal thickness (mm) | 2.98 (0.20) | 2.87 (0.29) | **.016** | 3.19 (0.22) | 2.95 (0.28) | **.007** |
| Parahippocampal thickness (mm) | 2.66 (0.20) | 2.65 (0.20) | .887 | 2.72 (0.20) | 2.58 (0.27) | .090 |
| eTIV (mm³) | 1532569.66 (142313.04) | 1540542.38 (138703.37) | .717 | 1468631.79 (232097.01) | 1462906.91 (176811.19) | .930 |
| ***Tau-PET SUVR*** |  |  |  |  |  |  |
| Amygdala tau-PET SUVR | 1.24 (0.11) | 1.35 (0.20) | **<.001** | 0.68 (0.13) | 1.06 (0.66) | **.036** |
| Entorhinal tau-PET SUVR | 1.04 (0.09) | 1.16 (0.18) | **<.001** | 0.85 (0.18) | 1.38 (0.64) | **.004** |
| Hippocampal tau-PET SUVR | 1.28 (0.11) | 1.32 (0.19) | .090 | 0.76 (0.11) | 1.00 (0.28) | **.004** |
| Parahippocampal tau-PET SUVR | 1.06 (0.08) | 1.12 (0.14) | **.001** | 0.82 (0.11) | 1.22 (0.43) | **.002** |

Note. Values are mean (SD) unless otherwise indicated; sex is reported as percentage female. PREVENT-AD p values compare CN A− with CN A+. UCLouvain p values compare CN A− with CN A+ using Welch two-sample t-tests for continuous variables and Fisher's exact tests for sex and APOE ε4 carriership. Education was measured in years in PREVENT-AD and coded categorically in UCLouvain (1 = low/no high school; 2 = medium/high school; 3 = high/university). CN = cognitively normal; A = amyloid; MoCA = Montreal Cognitive Assessment; MMSE = Mini-Mental State Examination; UPSIT = University of Pennsylvania Smell Identification Test; SUVR = standardized uptake value ratio; eTIV = estimated total intracranial volume. APOE ε4 carrier indicates the presence of at least one ε4 allele.
<sup>a</sup> PREVENT-AD MoCA score was missing for one participant; MRI was missing for 6 CN A− and 3 CN A+ participants; and tau-PET was missing for 2 CN A− participants.
<sup>b</sup> UCLouvain regional tau-PET was available for 13 CN A− and 17 CN A+ participants.

### 3.1 Group Comparisons

Mean odor identification scores were 87.1% (11.7%) and 81.8% (16.2%) in the PREVENT-AD CN A− and CN A+ groups, respectively. Corresponding values in UCLouvain were 75.0% (15.3%) and 63.5% (22.1%). After adjustment for cohort, age, sex, education, and APOE ε4-carrier status, CN A+ participants scored 4.64 percentage points lower than CN A− participants (adjusted difference, −4.64 percentage points; 95% CI, −8.60 to −0.68; *p* = .022) (Figure 1). The amyloid-status-by-cohort interaction was not significant (F[1,248] = 0.59; *p* = .445), indicating no evidence that the association differed between cohorts.

**Figure 1.**
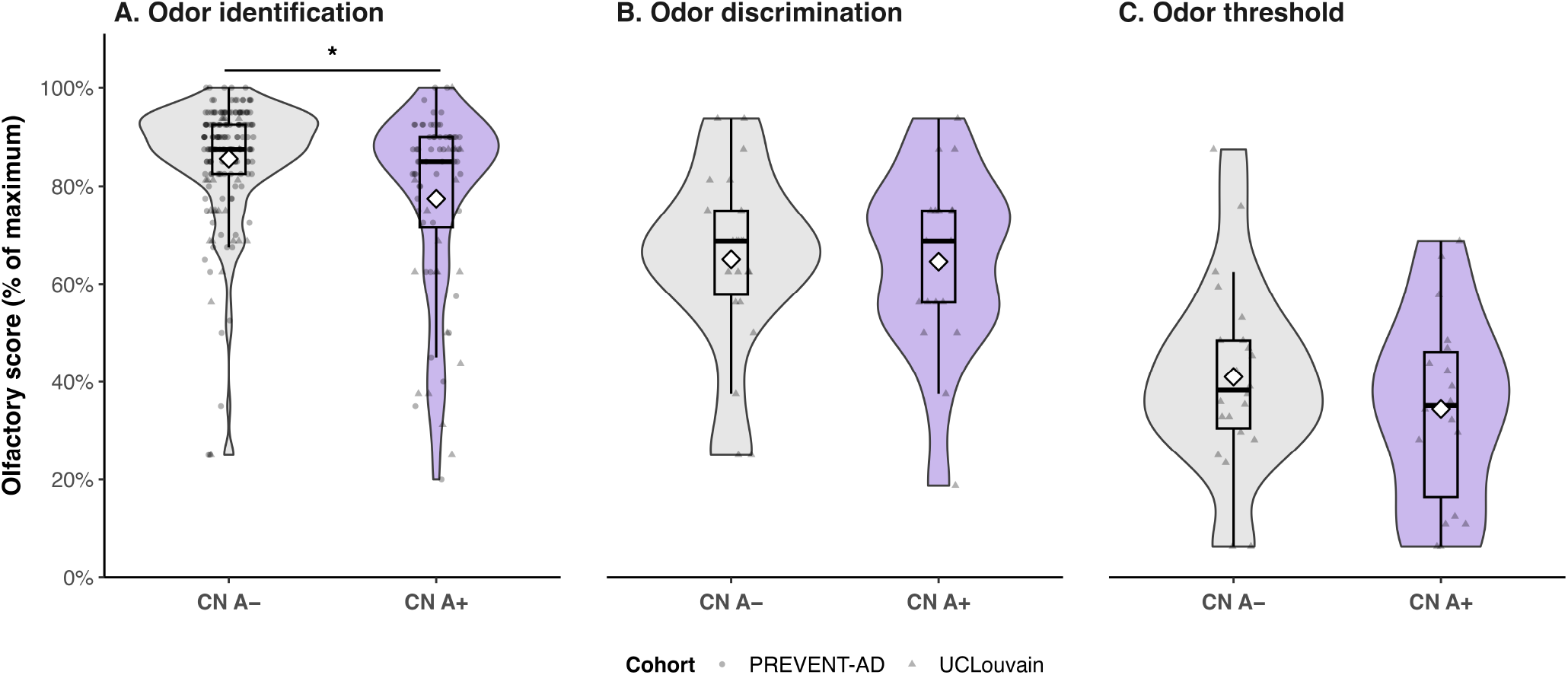
Olfactory scores by amyloid status. Distributions of odor identification scores in the combined PREVENT-AD and UCLouvain cohorts (A) and exploratory odor discrimination (B) and odor detection threshold (C) scores in UCLouvain. Scores are expressed as percentages of the maximum possible score. Violin plots show the distributions; boxes indicate medians and interquartile ranges, and diamonds indicate means. Points represent individual participants; circles denote PREVENT-AD and triangles denote UCLouvain. Gray represents CN A− participants, and purple represents CN A+ participants. The asterisk indicates the APOE ε4-adjusted CN A− versus CN A+ difference in odor identification (*p* = .022).

Exploratory UCLouvain analyses found no significant group differences in odor discrimination (adjusted difference, 4.62 percentage points; 95% CI, −7.45 to 16.69; *p* = .442) or odor detection threshold scores (−1.27 percentage points; 95% CI, −13.25 to 10.71; *p* = .831).

### 3.2 Neuroimaging Biomarker Associations

The median absolute interval between olfactory testing and tau-PET imaging was 17.0 months (IQR, 7.0– 55.5) in PREVENT-AD and 10.6 months (IQR, 7.7–22.2) in UCLouvain. For MRI imaging, the intervals were 14.5 months (IQR, 0–60) in PREVENT-AD and 10.7 months in UCLouvain (IQR, 8.1–22.5). First, in the combined-cohort models without regional interactions, no common MTL tau-PET or morphometric association survived FDR correction (Supplementary Table 1). Tau-PET associations differed between cohorts for the parahippocampal cortex (MTL FDR-adjusted *p* = .003), amygdala (MTL FDR-adjusted *p* = .030), and entorhinal cortex (MTL FDR-adjusted *p* = .044) (Table 2). Follow-up estimates showed that in UCLouvain, higher tau burden was associated with lower odor identification scores in the parahippocampal cortex (B = −12.81; 95% CI, −20.42 to −5.20; MTL FDR-adjusted *p* = .004) and amygdala (B = −6.59; 95% CI, −12.08 to −1.10; MTL FDR-adjusted *p* = .038). The UCLouvain entorhinal association did not survive FDR correction, and no corresponding MTL tau-PET associations were observed in PREVENT-AD (Table 2; Figure 2A). No MTL morphometric association or morphometry-by-cohort interaction survived FDR correction. Complete atlas-wide results are reported in Supplementary Table 2.

**Table 2.** Adjusted MTL biomarker-by-cohort interactions and follow-up cohort-specific associations.

| **Region** | **Cohort difference**<br>**B (95% CI)** | **FDR p** | **PREVENT-AD**<br>**B (95% CI)** | **FDR p** | **UCLouvain**<br>**B (95% CI)** | **FDR p** |
|---|---|---|---|---|---|---|
| Tau PET |  |  |  |  |  |  |
| Amygdala | −7.41 (−13.38 to −1.44) | **.030** | 0.82 (−1.42 to 3.06) | .471 | −6.59 (−12.08 to −1.10) | **.038** |
| Hippocampus | −5.95 (−15.76 to 3.86) | .234 | 1.42 (−1.42 to 4.27) | .434 | −4.52 (−13.90 to 4.85) | .343 |
| Entorhinal cortex | −11.55 (−22.15 to −0.96) | **.044** | 1.27 (−1.08 to 3.63) | .434 | −10.28 (−20.54 to −0.02) | .066 |
| Parahippocampal cortex | −14.49 (−22.74 to −6.24) | **.003** | 1.68 (−0.96 to 4.33) | .434 | −12.81 (−20.42 to −5.20) | **.004** |
| Morphometry |  |  |  |  |  |  |
| Amygdala | 1.88 (−4.19 to 7.94) | .543 | 2.37 (−0.91 to 5.65) | .624 | 4.25 (−1.89 to 10.38) | .174 |
| Hippocampus | 3.48 (−1.60 to 8.55) | .238 | 0.85 (−2.29 to 3.98) | .792 | 4.33 (−0.02 to 8.67) | .102 |
| Entorhinal cortex | 4.28 (−1.64 to 10.21) | .238 | 1.33 (−1.66 to 4.32) | .763 | 5.61 (0.51 to 10.71) | .102 |
| Parahippocampal cortex | 4.69 (−1.16 to 10.54) | .238 | −0.08 (−2.24 to 2.09) | .945 | 4.61 (−0.78 to 10.01) | .124 |

Note. B is the adjusted percentage-point change in odor-identification score per 1 within-cohort SD increase in the biomarker. Cohort difference is UCLouvain minus PREVENT-AD and represents the regional biomarker-by-cohort interaction. PREVENT-AD and UCLouvain slopes are follow-up estimates from the same interaction model. Models adjusted for cohort, amyloid status, age, sex, cohort-specific education, and APOE ε4-carrier status. Regional tau-PET models additionally included global cortical tau burden and its interactions with cohort and amyloid status; subcortical-volume models additionally included eTIV. Confidence intervals use HC1 robust standard errors. FDR p-values are Benjamini–Hochberg corrected across the four prespecified MTL regions separately by modality, cohort, and hypothesis.

**Figure 2.**
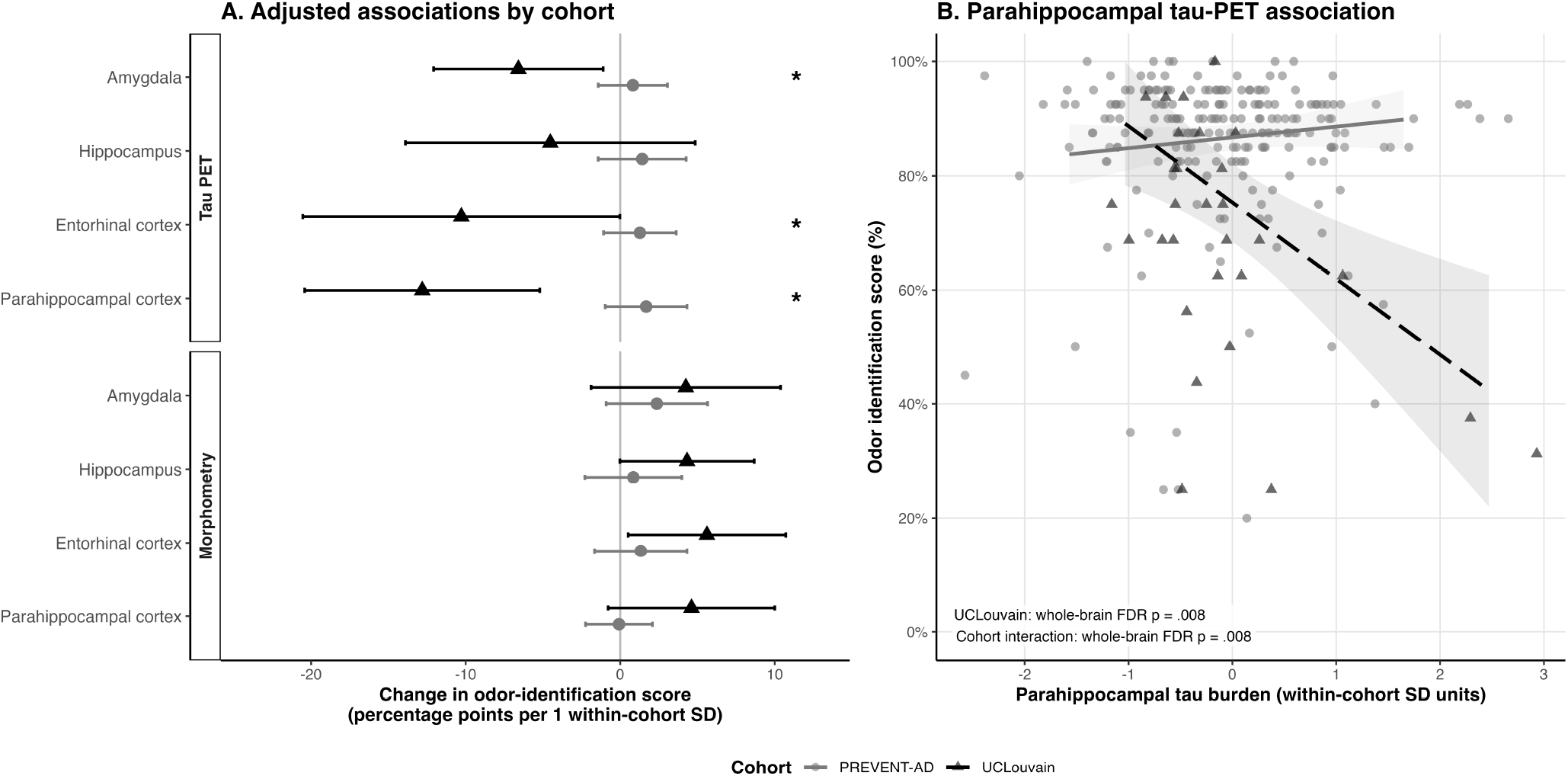
Regional neuroimaging biomarkers and odor identification performance. (A) Adjusted associations of medial temporal lobe tau-PET and morphometric measures with odor identification in each cohort. Points and horizontal lines represent adjusted coefficients and 95% confidence intervals, respectively. Asterisks indicate biomarker-by-cohort interactions that survived FDR correction across the four prespecified medial temporal lobe regions. (B) Sensitivity analysis of the parahippocampal tau-PET association after excluding regional or global cortical tau values exceeding ±3 within-cohort SDs. Points represent individual participants, fitted lines represent fully adjusted model predictions, and shaded areas represent 95% confidence intervals. Gray circles and solid lines indicate PREVENT-AD; black triangles and dashed lines indicate UCLouvain. All models were adjusted for age, sex, amyloid status, cohort-specific education, and APOE ε4-carrier status. Tau-PET models additionally included global cortical tau and its interactions with cohort and amyloid status; subcortical-volume models additionally included estimated total intracranial volume. The FDR-adjusted *p*-values displayed in panel B account for testing across all 42 atlas-wide regions.

#### 3.2.2 Amyloid status moderation

Amyloid status did not significantly moderate the associations of regional tau-PET or morphometry with odor identification across the prespecified MTL ROIs (Table 3; Supplementary Table 3). An exploratory hippocampal tau-PET-by-cohort-by-amyloid interaction was observed in the primary MTL analysis. However, it did not survive whole-brain FDR correction (Supplementary Table 4).

**Table 3.** Adjusted MTL amyloid-specific associations and interaction tests.

| Region | CN A−<br>B (95% CI) | CN A+<br>B (95% CI) | Amyloid interaction<br>B (95% CI) | FDR p | Cohort × amyloid interaction<br>B (95% CI) | FDR p |
|---|---|---|---|---|---|---|
| Tau PET |  |  |  |  |  |  |
| Amygdala | 3.14 (0.28 to 5.99) | −1.38 (−4.41 to 1.65) | −4.52 (−8.48 to −0.56) | .102 | −24.19 (−54.49 to 6.11) | .156 |
| Hippocampus | 2.44 (0.28 to 4.60) | −0.28 (−4.96 to 4.41) | −2.72 (−7.89 to 2.46) | .429 | −19.73 (−32.92 to −6.54) | **.014** |
| Entorhinal cortex | 1.96 (−0.99 to 4.92) | 0.13 (−3.15 to 3.41) | −1.84 (−6.03 to 2.36) | .429 | −10.67 (−23.22 to 1.89) | .156 |
| Parahippocampal cortex | 2.02 (−1.29 to 5.34) | −0.13 (−4.60 to 4.35) | −2.15 (−7.51 to 3.20) | .429 | −6.85 (−24.12 to 10.42) | .435 |
| Morphometry |  |  |  |  |  |  |
| Amygdala | 0.54 (−1.91 to 2.99) | 5.64 (0.55 to 10.73) | 5.10 (0.16 to 10.04) | .058 | −0.85 (−13.99 to 12.28) | .898 |
| Hippocampus | −0.31 (−2.67 to 2.05) | 4.23 (−1.03 to 9.50) | 4.55 (−0.93 to 10.02) | .103 | −9.69 (−23.94 to 4.56) | .726 |
| Entorhinal cortex | −0.40 (−2.28 to 1.47) | 4.79 (0.43 to 9.15) | 5.19 (0.38 to 10.00) | .058 | −3.45 (−15.47 to 8.57) | .780 |
| Parahippocampal cortex | −1.29 (−2.99 to 0.41) | 4.86 (−0.34 to 10.07) | 6.15 (0.56 to 11.74) | .058 | 2.90 (−7.53 to 13.32) | .780 |

**Note.** CN A− and CN A+ columns are adjusted biomarker slopes from the biomarker-by-amyloid model. Amyloid interaction is CN A+ minus CN A−. Cohort × amyloid interaction is the biomarker-by-cohort-by-amyloid coefficient and is exploratory. Models adjusted for cohort, age, sex, cohort-specific education, and APOE ε4-carrier status. Tau-PET models additionally included global cortical tau burden and its interactions with cohort and amyloid status; subcortical-volume models additionally included eTIV. FDR p-values are corrected across the four prespecified MTL regions separately by modality and hypothesis.

Among UCLouvain participants, visually assessed tau-positive (T+; n = 11) participants had lower mean odor identification scores than tau-negative (T−; n = 20) participants (56.2% vs 73.4%). After adjustment for amyloid status, age, sex, education, and APOE ε4-carrier status, the difference was not statistically significant (B = −13.90 percentage points; 95% CI, −30.56 to 2.76; *p* = .098; Supplementary Table 5).

#### 3.2.3 Sensitivity analysis

After excluding region-specific extreme imaging values, tau-PET models included 209–213 PREVENT-AD participants and 28–29 UCLouvain participants, depending on the region. Among the prespecified MTL ROIs, the parahippocampal cortex was the only region that remained significant after both extreme-value exclusion and whole-brain correction. Higher parahippocampal tau was associated with lower odor identification scores in UCLouvain (B = −13.36; 95% CI, −21.20 to −5.52; whole-brain FDR-adjusted *p* = .008), whereas no corresponding association was observed in PREVENT-AD (B = 1.89; 95% CI, −1.04 to 4.81). The association was more negative in UCLouvain than in PREVENT-AD (interaction B = −15.25; 95% CI, −23.82 to −6.68; whole-brain FDR-adjusted *p* = .008; Figure 2B).

The UCLouvain parahippocampal association remained significant when the olfaction-to-PET interval was restricted to 24 months or less (B = −14.91; 95% CI, −22.25 to −7.56; MTL FDR-adjusted *p* < .001) and 12 months or less (B = −13.90; 95% CI, −22.37 to −5.44; MTL FDR-adjusted *p* = .006). Corresponding cohort interactions remained significant at 24 months (MTL FDR-adjusted *p* = .003) and 12 months (MTL FDR-adjusted *p* = .034), although the 12-month UCLouvain analysis included only 17 participants (Supplementary Table 6). No morphometric association, regional biomarker-by-amyloid interaction, or regional biomarker-by-cohort-by-amyloid interaction survived whole-brain FDR correction after extreme-value exclusion. Complete extreme-value sensitivity results are presented in Supplementary Tables 7–11.

Overall, neuroimaging analyses identified a robust association between higher parahippocampal tau burden and lower odor identification in UCLouvain, whereas no corresponding association was observed in PREVENT-AD or for regional morphometry.

### 3.3 MCI Incidence

The analysis included 250 participants and 72 incident MCI events (28.8%). The mean follow-up time from olfactory testing to conversion or censoring was 5.63 years (SD = 1.48) in PREVENT-AD, and 3.45 years (SD = 3.78) in the UCLouvain cohort.

In the Cox model adjusted for amyloid status, age, sex, education, APOE ε4-carrier status, and baseline global cognition, each 10-percentage-point higher odor identification score was associated with a 16% lower hazard of incident MCI (HR, 0.84; 95% CI, 0.72–0.98; *p* = .027) (Figure 3). APOE ε4 carriership was not independently associated with incident MCI (HR, 1.14; 95% CI, 0.67–1.93; *p* = .621). There was no significant evidence that the odor-identification association differed by amyloid status (likelihood-ratio χ^2^[1] = 1.93; p = .164) or cohort (χ^2^[1] = 2.75; *p* = .097).

**Figure 3.**
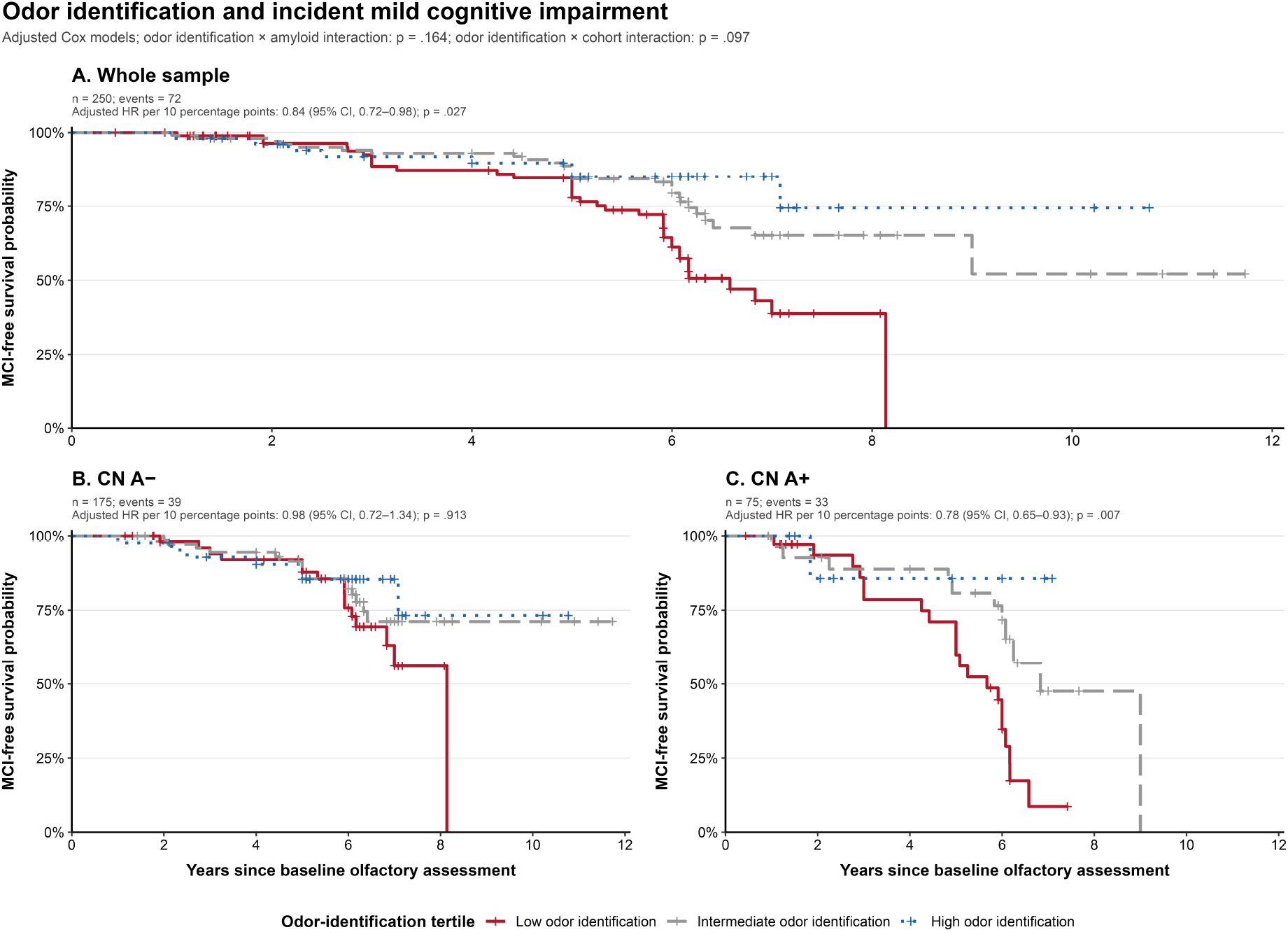
Odor identification and incident mild cognitive impairment. Kaplan–Meier curves show MCI-free survival according to cohort-specific tertiles of odor identification performance in (A) the complete sample, (B) CN A− participants, and (C) CN A+ participants. Curves are descriptive; crosses indicate censoring. Reported hazard ratios model odor identification continuously and represent the change in MCI hazard per 10-percentage-point higher score. Cox models were adjusted for amyloid status, age, sex, cohort-specific education, APOE ε4-carrier status, and baseline global cognition measured using the MoCA in PREVENT-AD and the MMSE in UCLouvain, with baseline hazards stratified by cohort. Neither the odor identification-by-amyloid interaction (likelihood-ratio p = .164) nor the odor identification-by-cohort interaction (likelihood-ratio p = .097) was significant.

## 4. Discussion

In this study, CN A+ participants showed lower odor identification compared with CN A− participants after adjustment for cohort, age, sex, education, and APOE ε4-carrier status. Exploratory UCLouvain analyses showed that these effects were not observed for odor discrimination or detection threshold. These findings suggest a specific olfactory impairment profile in which odor identification declines, whereas odor discrimination and detection threshold remain intact at the preclinical stage of AD ^29^. This selectivity is consistent with meta-analytic evidence showing greater odor identification impairment at the MCI ^45^ and dementia stages ^46^ and may reflect AD-related disruption of higher-order olfactory processes, including semantic access, memory, and MTL-dependent odor– object representations ^21,47,48^.

Neuroimaging analyses showed important cohort heterogeneity. In sensitivity analyses excluding extreme neuroimaging values and applying whole-brain FDR correction, higher tau-PET burden in the parahippocampal cortex was associated with lower odor identification in UCLouvain but not in PREVENT-AD. This regional finding is consistent with prior PET evidence linking poorer odor identification to greater parahippocampal tau ^27^ and with longitudinal evidence that odor identification deficits track tau accumulation and propagation from medial temporal structures through the olfactory network ^26^. The parahippocampal association, and difference across cohorts, were also retained when analyses were restricted to olfaction-to-PET intervals of 12 or 24 months. By contrast, neither MTL morphometry nor amyloid-status moderation survived FDR correction nor sensitivity analyses. Thus, the imaging findings support a tau–olfaction association in UCLouvain, but not a cohort-general effect.

This discrepancy may be partly explained by the use of different tau-PET tracers. PREVENT-AD used [^18^F]flortaucipir, a first-generation tau tracer, whereas UCLouvain used [^18^F]MK-6240, a second-generation tracer. Although both tracers detect AD-like tau pathology, they differ in binding characteristics and off-target signal. Direct head-to-head comparisons indicate that flortaucipir shows more frequent choroid plexus and higher striatal off-target binding, whereas MK-6240 more commonly shows pial/meningeal off-target signal ^49^. More recently, a large multicenter, within-participant comparison showed that [^18^F]MK-6240 detected MTL tau positivity more frequently than flortaucipir, particularly among cognitively unimpaired A+ individuals, and reached tau positivity at lower amyloid burden ^50^. In preclinical AD, where tau burden is subtle and spatially restricted, the stronger tau-PET/odor identification associations observed in UCLouvain, despite its smaller sample size, may therefore partly reflect greater sensitivity of [^18^F]MK-6240 to early MTL tau signal.

Longitudinal analyses showed that lower baseline odor identification scores predicted progression to MCI independently of baseline global cognition, age, sex, education, and APOE ε4-carrier status. Each 10-percentage-point higher score was associated with a 16% lower hazard of incident MCI. Formal interaction tests provided no evidence that this association differed by amyloid status or cohort. Clinically, odor identification testing could serve as a low-cost, scalable first-stage screen for AD-related cognitive decline. Because poor olfactory function is nonspecific to AD, it should prompt evaluation for other causes of olfactory dysfunction and, when appropriate, confirmatory biomarker assessment. Established tests such as the UPSIT ^36^ and Sniffin’ Sticks ^37^, as well as olfactory digital tools such as the AROMHA Brain Health Test ^51^, could support scalable risk stratification.

This study has some limitations. The PREVENT-AD and UCLouvain cohorts differed in recruitment strategies, olfactory tests, and tau-PET tracers, which may have contributed to cohort-specific findings. Sample sizes also varied, with larger power in PREVENT-AD and more limited sensitivity in UCLouvain. In addition, although imaging protocols were processed with similar standard pipelines, scanner and acquisition differences could introduce technical variability. Finally, most of our analyses were cross-sectional, limiting our ability to draw conclusions about the temporal sequence between olfactory decline, structural atrophy, and tau aggregation.

To conclude, odor identification was lower in CN A+ older adults and was associated with incident MCI after adjustment for APOE ε4-carrier status and baseline cognitive function. The association with parahippocampal cortex tau burden differed across cohorts and was present in UCLouvain ([^18^F]MK-6240) but absent in PREVENT-AD ([^18^F]flortaucipir). Odor identification may therefore provide a low-cost indicator of clinical risk, while its neuroimaging correlates require replication using harmonized tracers and acquisition protocols.

## Data Availability

Data used in preparation of this article were obtained from the PREVENT-AD Registered repository available at https://registeredpreventad.loris.ca. UCLouvain de-identified data are available upon request to the authors, subject to restricted access through the corresponding author.

## Acknowledgements

The authors thank the participants for their important contributions. ChatGPT was used for English-language editing and assistance with coding. ChatGPT was not used for study design, data collection, statistical analysis, interpretation of the results, or generation of research data. The authors reviewed and edited all AI-assisted content.

## Funding

The PREVENT-AD program was supported by the Canadian Institutes of Health Research (CIHR), the Canada First Research Excellence Fund (CFREF) awarded to Healthy Brains, Healthy Lives (HBHL), the Fonds de Recherche du Québec – Santé (FRQS), the Douglas Mental Health University Institute Foundation, and by philanthropic contributions to the StoP-AD Centre at the Douglas Research Centre. Additional support for PREVENT-AD infrastructure, participant follow-up, and biomarker acquisition was provided through CIHR grants awarded to the principal investigators of the program. The UCLouvain cohort (PI: B.H.) was supported by the Belgian Fund for Scientific Research (FNRS), the Belgian StopAlzheimer Foundation (SAO-FRA), and the WEL Research Institute. Data collection was additionally supported by institutional research funds from the Institute of Neuroscience (IoNS) at UCLouvain and the Cliniques Universitaires Saint-Luc. Tau-PET imaging (MK-6240) was made possible through collaborations with Lantheus, KULeuven, and the WEL Research Institute. B. Jobin is supported by scholarships from the Canadian Institutes of Health Research (CIHR) and the Université du Québec à Trois-Rivières. B. Hanseeuw is a clinical scientist (CCL) of the Belgian Fonds de la Recherche Scientifique (FNRS). Y. Salman is FNRS (FRIA) PhD fellow.

## Conflict of Interest Statement

B. Hanseeuw has served as a consultant for Biogen, Eisai, Eli Lilly, and Roche, outside the submitted work.

## Consent Statement

The PREVENT-AD study was approved by the institutional review boards of McGill University and the Douglas Mental Health University Institute. The UCLouvain study was approved by the Ethics Committee of UCLouvain (UCL-2016–121, Date: 13/05/2019; Eudra-CT number: 2018–0034/73–94; 2010/17May/148). All participants provided written informed consent prior to participation, in accordance with the Declaration of Helsinki.

## Authors Contributions

B. Jobin: Conceptualization, Formal analysis, Investigation, Methodology, Project administration, Visualization, Writing - original draft.

Y. Salman: Conceptualization, Investigation, Methodology, Neuroimaging Processing, Project administration, Writing - review and editing.

K. Bouchoucha: Data acquisition, Writing - review and editing.

L. Huyghe: Data acquisition, Writing - review and editing.

L. Quenon: Data acquisition, Writing - review and editing.

L. Colmant: Data acquisition, Neuroimaging Processing, Writing - review and editing.

T. Gérard: Data acquisition, Writing - review and editing.

V. Malotaux: Data acquisition, Neuroimaging Processing, Writing - review and editing.

L. Dricot: Ressources, Writing - review and editing.

P. Dulieu: Data acquisition, Resources, Writing - review and editing.

A. Ivanoiu: Writing - review and editing.

R. Lhommel: Data acquisition, Resources, Writing - review and editing.

A. Decottignies: Funding acquisition, Writing – review & editing.

C. Huart: Funding acquisition, Methodology, Supervision, Writing – review & editing.

B. Hanseeuw: Funding acquisition, Methodology, Supervision, Writing – review & editing.

PREVENT-AD Research Group: Conceptualization, Project administration, Methodology, Funding acquisition.

## Notes

### Author Declarations

The institutional review boards of McGill University and the Douglas Mental Health University Institute gave ethical approval for the PREVENT-AD study. The Ethics Committee of UCLouvain (Universite catholique de Louvain) gave ethical approval for the UCLouvain study (UCL-2016-121, Date: 13/05/2019; Eudra-CT number: 2018-0034/73-94; 2010/17May/148). All participants provided written informed consent prior to participation, in accordance with the Declaration of Helsinki.

## References

1. Frisoni GB, Hansson O, Nichols E, et al. New landscape of the diagnosis of Alzheimer’s disease. The Lancet. Published online September 2025:S0140673625012942. doi:10.1016/S0140-6736(25)01294-2

2. Jack CR, Bennett DA, Blennow K, et al. NIA-AA Research Framework: Toward a biological definition of Alzheimer’s disease. Alzheimer’s & Dementia. 2018;14(4):535–562. doi:10.1016/j.jalz.2018.02.018

3. De Strooper B, Karran E. The Cellular Phase of Alzheimer’s Disease. Cell. 2016;164(4):603–615. doi:10.1016/j.cell.2015.12.056

4. Harrison TM, La Joie R, Maass A, et al. Longitudinal tau accumulation and atrophy in aging and alzheimer disease. Annals of Neurology. 2019;85(2):229–240. doi:10.1002/ana.25406

5. Jack CR, Andrews JS, Beach TG, et al. Revised criteria for diagnosis and staging of Alzheimer’s disease: Alzheimer’s Association Workgroup. Alzheimer’s & Dementia. Published online June 27, 2024:alz.13859. doi:10.1002/alz.13859

6. Jia J, Ning Y, Chen M, et al. Biomarker Changes during 20 Years Preceding Alzheimer’s Disease. N Engl J Med. 2024;390(8):712–722. doi:10.1056/NEJMoa2310168

7. Sperling RA, Aisen PS, Beckett LA, et al. Toward defining the preclinical stages of Alzheimer’s disease: Recommendations from the National Institute on Aging-Alzheimer’s Association workgroups on diagnostic guidelines for Alzheimer’s disease. Alzheimer’s & Dementia. 2011;7(3):280–292. doi:10.1016/j.jalz.2011.03.003

8. Mintun MA, Lo AC, Evans CD, et al. Donanemab in Early Alzheimer’s Disease. n engl j med. 2021;384(18):1691–1704. doi:10.1056/NEJMoa2100708

9. Sims JR, Zimmer JA, Evans CD, et al. Donanemab in Early Symptomatic Alzheimer Disease: The TRAILBLAZER-ALZ 2 Randomized Clinical Trial. JAMA. 2023;330(6):512. doi:10.1001/jama.2023.13239

10. van Dyck CH, Swanson CJ, Aisen P, et al. Lecanemab in Early Alzheimer’s Disease. n engl j med. 2023;388(1):9–21. doi:10.1056/NEJMoa2212948

11. Palmqvist S, Whitson HE, Allen LA, et al. Alzheimer’s Association Clinical Practice Guideline on the use of blood ‐based biomarkers in the diagnostic workup of suspected Alzheimer’s disease within specialized care settings. Alzheimer’s & Dementia. 2025;21(7):e70535. doi:10.1002/alz.70535

12. Braak H, Braak E. Neuropathological stageing of Alzheimer-related changes. Acta Neuropathol. 1991;82(4):239–259. doi:10.1007/BF00308809

13. Kovács T, Cairns NJ, Lantos PL. Olfactory centres in Alzheimer’s disease: olfactory bulb is involved in early Braak’s stages: Neuroreport. 2001;12(2):285–288. doi:10.1097/00001756-200102120-00021

14. Baumeister H, Gellersen HM, Polk SE, et al. Disease stage-specific atrophy markers in Alzheimer’s disease. Neurology. Preprint posted online March 14, 2025. doi:10.1101/2025.03.13.25323904

15. Jobin B, Boller B, Frasnelli J. Smaller grey matter volume in the central olfactory system in mild cognitive impairment. Experimental Gerontology. 2023;183:112325. doi:10.1016/j.exger.2023.112325

16. Planche V, Manjon JV, Mansencal B, et al. Structural progression of Alzheimer’s disease over decades: the MRI staging scheme. Brain Communications. 2022;4(3):fcac109. doi:10.1093/braincomms/fcac109

17. Salman Y, Gérard T, Huyghe L, et al. Amygdala atrophies in specific subnuclei in preclinical Alzheimer’s disease. Alzheimer’s & Dementia. Published online September 10, 2024:alz.14235. doi:10.1002/alz.14235

18. Jobin B, Zahal R, Bussières EL, Frasnelli J, Boller B. Olfactory Identification in Subjective Cognitive Decline: A Meta-Analysis. Pike K, ed. JAD. 2021;79(4):1497–1507. doi:10.3233/JAD-201022

19. Jobin B, Zigrand C, Frasnelli J, Boller B, Albers MW. Lower odor identification in subjective cognitive decline: a meta‐analysis. Alz & Dem Diag Ass & Dis Mo. 2025;17(3):e70168. doi:10.1002/dad2.70168

20. Jobin B, Phillips NA, Frasnelli J, Boller B, CIMA-Q group. The predictive role of olfactory identification on episodic memory and mild cognitive impairment: Results from the CIMA-Q cohort. Journal of Alzheimer’s Disease. Published online 2025:13872877251378386.

21. Jobin B, Roy-Côté F, Frasnelli J, Boller B. Olfaction and Declarative Memory in Aging: A Meta-analysis. Chemical Senses. 2023;48:bjad045. doi:10.1093/chemse/bjad045

22. Devanand DP, Lee S, Manly J, et al. Olfactory deficits predict cognitive decline and Alzheimer dementia in an urban community. Neurology. 2015;84(2):182–189. doi:10.1212/WNL.0000000000001132

23. Dhilla Albers A, Asafu-Adjei J, Delaney MK, et al. Episodic memory of odors stratifies Alzheimer biomarkers in normal elderly: POEM: Odor Memory Biomarker in Normal Elderly. Ann Neurol. 2016;80(6):846–857. doi:10.1002/ana.24792

24. Murphy C, Jernigan TL, Fennema-Notestine C. Left hippocampal volume loss in Alzheimer’s disease is reflected in performance on odor identification: A structural MRI study. J Int Neuropsychol Soc. 2003;9(3):459–471. doi:10.1017/S1355617703930116

25. Papadatos Z, Phillips NA. Olfactory function reflects episodic memory performance and atrophy in the medial temporal lobe in individuals at risk for Alzheimer’s disease. Neurobiology of Aging. 2023;128:33–42. doi:10.1016/j.neurobiolaging.2023.04.001

26. Diez I, Ortiz-Terán L, Ng TSC, et al. Tau propagation in the brain olfactory circuits is associated with smell perception changes in aging. Nat Commun. 2024;15(1):4809. doi:10.1038/s41467-024-48462-3

27. Risacher SL, Tallman EF, West JD, et al. Olfactory identification in subjective cognitive decline and mild cognitive impairment: Association with tau but not amyloid positron emission tomography. Alzheimer’s & Dementia: Diagnosis, Assessment & Disease Monitoring. 2017;9:57–66.

28. Klein J, Yan X, Johnson A, et al. Olfactory Impairment Is Related to Tau Pathology and Neuroinflammation in Alzheimer’s Disease. Gordon B, ed. JAD. 2021;80(3):1051–1065. doi:10.3233/JAD-201149

29. Bouchoucha K, Collin L, Coquette C, et al. Olfactory decline tracks central‐to‐peripheral spread of tau pathology in Alzheimer’s disease. Alzheimer’s & Dementia. 2026;22(4):e71402. doi:10.1002/alz.71402

30. Nasreddine ZS, Phillips NA, Bédirian V, et al. The Montreal Cognitive Assessment, MoCA: A Brief Screening Tool For Mild Cognitive Impairment: MOCA: A BRIEF SCREENING TOOL FOR MCI. Journal of the American Geriatrics Society. 2005;53(4):695–699. doi:10.1111/j.1532-5415.2005.53221.x

31. Morris JC. The Clinical Dementia Rating (CDR) current version and scoring rules. Neurology. 1993;43(11):2412–2412.

32. Tremblay-Mercier J, Madjar C, Das S, et al. Open science datasets from PREVENT-AD, a longitudinal cohort of pre-symptomatic Alzheimer’s disease. NeuroImage: Clinical. 2021;31:102733. doi:10.1016/j.nicl.2021.102733

33. Villeneuve S, Poirier J, Breitner JCS, et al. The PREVENT‐AD cohort: Accelerating Alzheimer’s disease research and treatment in Canada and beyond. Alzheimer’s & Dementia. 2025;21(10):e70653. doi:10.1002/alz.70653

34. Amadoru S, Doré V, McLean CA, et al. Comparison of amyloid PET measured in Centiloid units with neuropathological findings in Alzheimer’s disease. Alz Res Therapy. 2020;12(1):22. doi:10.1186/s13195-020-00587-5

35. Ivanoiu A, Dricot L, Gilis N, et al. Classification of Non-Demented Patients Attending a Memory Clinic using the New Diagnostic Criteria for Alzheimer’s Disease with Disease-Related Biomarkers. JAD. 2014;43(3):835–847. doi:10.3233/JAD-140651

36. Doty RL, Shaman P, Kimmelman CP, Dann MS. University of Pennsylvania Smell Identification Test: a rapid quantitative olfactory function test for the clinic. The Laryngoscope. 1984;94(2):176–178.

37. Hummel T, Sekinger B, Wolf SR, Pauli E, Kobal G. ‘Sniffin’sticks’: olfactory performance assessed by the combined testing of odor identification, odor discrimination and olfactory threshold. Chemical senses. 1997;22(1):39–52.

38. Wolfensberger M. Sniffin’Sticks: a new olfactory test battery. Acta oto-laryngologica. 2000;120(2):303–306.

39. Haehner A, Mayer AM, Landis BN, et al. High test–retest reliability of the extended version of the “Sniffin’Sticks” test.Chemical senses. 2009;34(8):705–711.

40. Desikan RS, Ségonne F, Fischl B, et al. An automated labeling system for subdividing the human cerebral cortex on MRI scans into gyral based regions of interest. NeuroImage. 2006;31(3):968–980. doi:10.1016/j.neuroimage.2006.01.021

41. Malotaux V, Colmant L, Quenon L, et al. Suspecting Non-Alzheimer’s Pathologies and Mixed Pathologies: A Comparative Study Between Brain Metabolism and Tau Images. JAD. 2024;97(1):421–433. doi:10.3233/JAD-230696

42. Lundström JN, Boesveldt S, Albrecht J. Central Processing of the Chemical Senses: An Overview. ACS Chem Neurosci. 2011;2(1):5–16. doi:10.1021/cn1000843

43. Seubert J, Freiherr J, Djordjevic J, Lundström JN. Statistical localization of human olfactory cortex. Neuroimage. 2013;66:333–342.

44. Torske A, Koch K, Eickhoff S, Freiherr J. Localizing the human brain response to olfactory stimulation: A meta-analytic approach. Neuroscience & Biobehavioral Reviews. Published online December 2021:S0149763421005832. doi:10.1016/j.neubiorev.2021.12.035

45. Roalf DR, Moberg MJ, Turetsky BI, et al. A quantitative meta-analysis of olfactory dysfunction in mild cognitive impairment. J Neurol Neurosurg Psychiatry. 2017;88(3):226–232. doi:10.1136/jnnp-2016-314638

46. Rahayel S, Frasnelli J, Joubert S. The effect of Alzheimer’s disease and Parkinson’s disease on olfaction: a meta-analysis. Behavioural Brain Research. 2012;231(1):60–74. doi:10.1016/j.bbr.2012.02.047

47. Kjelvik G, Evensmoen HR, Hummel T, et al. The Human Brain Representation of Odor Identification in Amnestic Mild Cognitive Impairment and Alzheimer’s Dementia of Mild Degree. Frontiers in Neurology. 2021;11:1779. doi:10.3389/fneur.2020.607566

48. Kjelvik G, Evensmoen HR, Brezova V, Håberg AK. The human brain representation of odor identification. Journal of Neurophysiology. 2012;108(2):645–657. doi:10.1152/jn.01036.2010

49. Gogola A, Minhas DS, Villemagne VL, et al. Direct Comparison of the Tau PET Tracers18 F-Flortaucipir and18 F-MK-6240 in Human Subjects. J Nucl Med. 2022;63(1):108–116. doi:10.2967/jnumed.120.254961

50. Povala G, Bellaver B, Lussier FZ, et al. Comparison of [18F]flortaucipir and [18F]MK6240 for the detection of tau pathology in Alzheimer’s disease (HEAD): a multicentre, prospective, cross-sectional, within-participant study. The Lancet. 2026;407(10544):2217–2226. doi:10.1016/S0140-6736(26)00417-4

51. Jobin B, Magdamo C, Delphus D, et al. The AROMHA brain health test is a remote olfactory assessment to screen for cognitive impairment. Scientific Reports. 2025;15(9290).

