## Supplementary Methods and Tables for "Olfactory Function in Preclinical Alzheimer’s Disease: A Multi-Cohort Study"

**Supplementary Neuroimaging Acquisition Methods**

#### *Neuroimaging Acquisition*

##### PREVENT-AD Cohort.

***MRI.*** MRI scans were acquired at the Cerebral Imaging Center of the Douglas Mental Health University Institute in Montréal. During Phase 1 (2011–2017), T1-weighted neuroimaging was performed using a 3T Siemens Tim Trio scanner with a standard 12-channel head coil. The protocol was harmonized with the ADNI initiative and included T1-weighted MPRAGE sequence. For participants recruited after this date, the protocol was updated to include quantitative T1 mapping (MP2RAGE), acquired with a 32-channel head coil. In Phase 2 (2019 onward), MRI acquisition resumed following an upgrade to a 3T Siemens Prisma Fit scanner equipped with a 32-channel head coil. Sequences included 1 $mm^3$ 3D T1-weighted MPRAGE. All MRI data underwent defacing and quality control examination.

***[$^{18}$F]Flortaucipir Tau-PET.*** Tau-PET was performed at the McConnell Brain Imaging Center (Montreal Neurological Institute) using a Siemens/CTI high-resolution research tomograph (HRRT), with a spatial resolution of approximately 2.4 mm at the center of the field of view. A first-generation tau tangles imaging agent [$^{18F}$]Flortaucipir (370 MBq) was administered, and four 5-minute frames were collected 80 to 100 minutes post-injection. Images were corrected for attenuation, scatter, and decay, and standardized uptake value ratio (SUVR) images were generated using the inferior cerebellar gray matter as reference region.

##### UCLouvain Cohort.

***MRI.***

MRI data were acquired at the Cliniques Universitaires Saint-Luc in Brussels. Before October 2019, 3D T1-weighted images were acquired on a 3T Achieva MRI scanner (Philips Healthcare) using a 32-channel phased-array head coil. One hundred and fifty sagittal slices were acquired with the following parameters:

TR/TE/FA = 9.1 ms/4.6 ms/8°; slice thickness = 1 mm; acquisition resolution = 0.81 × 0.95 mm², reconstructed to 0.75 × 0.75 mm²; FOV = 220 × 197 mm²; acquisition matrix = 296 × 247; SENSE factor = 1.5 (parallel imaging).

After the replacement of this scanner in October 2019, 3D T1-weighted images were acquired on a 3T SIGNA™ Premier® MRI system (GE Healthcare) using a 48-channel phased-array head coil. One hundred and ninety-six sagittal slices were acquired with the following parameters: TR/TE/FA = 7.2 ms/2.9 ms/11°; slice thickness = 1.2 mm; FOV = 270 × 270 mm²; acquisition matrix = 256 × 256; acquisition resolution = 1.055 × 1.055 mm²; ASSET factor = 1.75 (parallel imaging).

All MRI data underwent quality control examination.

***[$^{18}$F]MK6240 Tau-PET.*** [$^{18}$F]MK6240 (Lantheus Inc.) is an investigational drug under study as a second-generation cerebral tau tangles imaging agent. Radiosynthesis was conducted at KULeuven and delivered in less than an hour. Ninety minutes after intravenous administration of [$^{18}$F]MK6240 (target activity 185 ± 5 MBq) a 30-minute dynamic list-mode acquisition was performed on a Philips Vereos digital PET-CT (Philips Healthcare). Images were reconstructed using the manufacturer's reconstruction algorithm, which includes attenuation, scatter, decay correction, and time-of-flight information. Point spread function (PSF) and 1mm reslicing were also computed using the manufacturer's algorithm to obtain a better resolution recovery. The cerebellar gray matter was used as a reference to generate MK6240 SUVR.

#### Supplementary Table 1. Common MTL biomarker associations across cohorts

| Modality | Region | B (95% CI) | FDR p |
|---|---|---|---|
| Tau PET | Amygdala | 0.12 (−2.31 to 2.56) | .920 |
| Tau PET | Hippocampus | 0.99 (−1.83 to 3.82) | .678 |
| Tau PET | Entorhinal cortex | 0.81 (−1.60 to 3.21) | .678 |
| Tau PET | Parahippocampal cortex | 0.98 (−1.86 to 3.82) | .678 |
| Morphometry | Amygdala | 2.64 (−0.51 to 5.80) | .269 |
| Morphometry | Hippocampus | 1.43 (−1.33 to 4.19) | .412 |
| Morphometry | Entorhinal cortex | 1.95 (−0.61 to 4.51) | .269 |
| Morphometry | Parahippocampal cortex | 0.68 (−1.44 to 2.80) | .528 |

Note. Common-association models estimated one regional biomarker slope across cohorts without a regional biomarker-by-cohort interaction. B is the adjusted percentage-point change in odor-identification score per 1 within-cohort SD higher biomarker value. Models adjusted for cohort, amyloid status, age, sex, and cohort-specific education and APOE ε4-carrier status. Tau-PET models also included global cortical tau and its interactions with cohort and amyloid status; subcortical-volume models included eTIV. FDR p-values are corrected across the four prespecified MTL regions separately by modality.

#### Supplementary Table 2. Whole-brain biomarker-by-cohort interactions and follow-up cohort-specific associations

| **Region** | **Cohort difference B (95% CI)** | FDR p | **PREVENT-AD B (95% CI)** | FDR p | **UCLouvain B (95% CI)** | FDR p |
|---|---|---|---|---|---|---|
| Tau PET | | | | | | |
| Banks of superior temporal sulcus | −1.36 (−17.35 to 14.63) | .989 | −0.22 (−3.12 to 2.68) | .957 | −1.58 (−17.69 to 14.53) | .983 |
| Caudal anterior cingulate | 4.36 (−9.03 to 17.76) | .843 | 0.22 (−2.66 to 3.11) | .957 | 4.58 (−8.66 to 17.83) | .771 |
| Caudal middle frontal | 9.42 (−9.97 to 28.80) | .594 | 1.86 (−2.45 to 6.17) | .774 | 11.27 (−7.63 to 30.18) | .461 |
| Cuneus | −0.18 (−8.01 to 7.65) | .989 | 2.20 (−0.45 to 4.85) | .530 | 2.02 (−5.14 to 9.19) | .838 |
| Entorhinal cortex | −11.55 (−22.15 to −0.96) | .143 | 1.27 (−1.08 to 3.63) | .774 | −10.28 (−20.54 to −0.02) | .179 |
| Frontal pole | 5.81 (−3.60 to 15.21) | .491 | −0.17 (−2.59 to 2.25) | .957 | 5.64 (−3.42 to 14.69) | .461 |
| Fusiform gyrus | −13.75 (−20.58 to −6.92) | **.002** | −0.88 (−3.46 to 1.69) | .774 | −14.64 (−20.83 to −8.45) | **<.001** |
| Inferior parietal | −12.89 (−38.05 to 12.27) | .579 | −3.57 (−8.14 to 1.01) | .530 | −16.46 (−41.73 to 8.82) | .444 |
| Inferior temporal | −18.08 (−35.88 to −0.29) | .177 | −1.96 (−5.17 to 1.24) | .739 | −20.05 (−37.65 to −2.44) | .135 |
| Insula | −5.53 (−38.33 to 27.28) | .989 | 3.01 (−0.08 to 6.11) | .396 | −2.51 (−35.22 to 30.20) | .983 |
| Isthmus cingulate | −0.88 (−22.35 to 20.60) | .989 | 1.11 (−1.13 to 3.36) | .774 | 0.24 (−21.27 to 21.75) | .983 |
| Lateral occipital | −7.74 (−10.72 to −4.76) | **<.001** | 0.89 (−0.88 to 2.66) | .774 | −6.85 (−9.41 to −4.29) | **<.001** |
| Lateral orbitofrontal | 3.20 (−24.54 to 30.94) | .989 | −0.61 (−3.99 to 2.77) | .922 | 2.59 (−24.93 to 30.11) | .983 |
| Lingual gyrus | −7.19 (−12.66 to −1.72) | .071 | 2.25 (0.12 to 4.39) | .342 | −4.94 (−9.88 to −0.00) | .179 |
| Medial orbitofrontal | 20.99 (−0.95 to 42.93) | .212 | 0.75 (−2.10 to 3.59) | .848 | 21.74 (−0.08 to 43.55) | .179 |
| Middle temporal | −2.33 (−16.99 to 12.32) | .989 | −0.00 (−2.98 to 2.97) | 1.000 | −2.33 (−16.98 to 12.31) | .983 |
| Paracentral lobule | 5.02 (−1.83 to 11.87) | .395 | 1.37 (−1.55 to 4.29) | .774 | 6.39 (−0.03 to 12.81) | .179 |
| Parahippocampal cortex | −14.49 (−22.74 to −6.24) | **.009** | 1.68 (−0.96 to 4.33) | .739 | −12.81 (−20.42 to −5.20) | **.011** |
| Pars opercularis | 13.77 (−7.74 to 35.29) | .487 | 1.40 (−2.21 to 5.02) | .774 | 15.18 (−6.03 to 36.38) | .395 |
| Pars orbitalis | 7.18 (−3.38 to 17.73) | .449 | −0.88 (−3.53 to 1.78) | .774 | 6.30 (−4.05 to 16.65) | .461 |
| Pars triangularis | 14.67 (5.05 to 24.28) | **.031** | 0.98 (−1.13 to 3.09) | .774 | 15.65 (6.09 to 25.21) | **.012** |
| Pericalcarine cortex | 0.81 (−5.44 to 7.05) | .989 | 1.73 (−0.63 to 4.09) | .570 | 2.54 (−3.28 to 8.35) | .631 |
| Postcentral gyrus | 8.48 (1.61 to 15.35) | .083 | −0.49 (−3.49 to 2.51) | .922 | 7.99 (1.53 to 14.45) | .109 |
| Posterior cingulate | −1.29 (−34.85 to 32.27) | .989 | 0.66 (−2.31 to 3.63) | .894 | −0.62 (−34.09 to 32.84) | .983 |
| Precentral gyrus | 15.06 (4.63 to 25.50) | **.041** | 2.70 (−0.66 to 6.05) | .530 | 17.76 (7.66 to 27.86) | **.009** |
| Precuneus | 4.03 (−3.47 to 11.52) | .579 | −0.13 (−4.59 to 4.32) | .976 | 3.89 (−2.05 to 9.84) | .444 |
| Rostral anterior cingulate | 10.14 (−2.45 to 22.72) | .342 | 0.29 (−2.00 to 2.57) | .957 | 10.42 (−2.05 to 22.89) | .265 |
| Rostral middle frontal | 13.46 (−1.26 to 28.18) | .235 | −1.05 (−3.81 to 1.70) | .774 | 12.41 (−1.96 to 26.77) | .252 |
| Superior frontal | 10.26 (−9.91 to 30.43) | .579 | −1.37 (−5.22 to 2.48) | .774 | 8.89 (−10.55 to 28.33) | .631 |
| Superior parietal | 0.93 (−6.84 to 8.70) | .989 | −1.56 (−5.25 to 2.13) | .774 | −0.63 (−7.67 to 6.40) | .983 |
| Superior temporal | −0.60 (−28.05 to 26.86) | .989 | 0.13 (−3.90 to 4.15) | .976 | −0.47 (−28.15 to 27.21) | .983 |
| Supramarginal gyrus | 19.75 (1.50 to 38.01) | .143 | −4.18 (−7.88 to −0.49) | .342 | 15.57 (−2.31 to 33.45) | .252 |
| Temporal pole | −5.58 (−23.77 to 12.61) | .849 | −2.44 (−5.12 to 0.25) | .449 | −8.02 (−25.84 to 9.80) | .631 |
| Transverse temporal | −1.40 (−9.05 to 6.25) | .989 | 2.61 (0.24 to 4.98) | .342 | 1.20 (−6.09 to 8.50) | .983 |
| Thalamus | 5.80 (−1.96 to 13.57) | .395 | 0.47 (−2.37 to 3.31) | .922 | 6.27 (−0.87 to 13.42) | .252 |
| Caudate | −0.82 (−14.65 to 13.01) | .989 | 1.85 (0.14 to 3.56) | .342 | 1.03 (−12.68 to 14.74) | .983 |
| Putamen | 1.87 (−11.53 to 15.26) | .989 | 0.70 (−1.81 to 3.21) | .847 | 2.56 (−10.60 to 15.73) | .982 |
| Pallidum | −0.10 (−14.09 to 13.90) | .989 | 0.25 (−2.18 to 2.69) | .957 | 0.15 (−13.69 to 14.00) | .983 |
| Hippocampus | −5.95 (−15.76 to 3.86) | .491 | 1.42 (−1.42 to 4.27) | .774 | −4.52 (−13.90 to 4.85) | .626 |
| Amygdala | −7.41 (−13.38 to −1.44) | .083 | 0.82 (−1.42 to 3.06) | .774 | −6.59 (−12.08 to −1.10) | .113 |
| Accumbens area | −8.53 (−29.39 to 12.33) | .708 | 2.08 (0.09 to 4.07) | .342 | −6.45 (−27.29 to 14.39) | .814 |
| Ventral diencephalon | −0.56 (−11.34 to 10.21) | .989 | 0.98 (−1.50 to 3.47) | .774 | 0.42 (−10.16 to 11.00) | .983 |
| Morphometry | | | | | | |
| Banks of superior temporal sulcus | 0.92 (−4.42 to 6.26) | .941 | 0.29 (−1.62 to 2.20) | .994 | 1.21 (−3.89 to 6.31) | .870 |
| Caudal anterior cingulate | 5.77 (1.47 to 10.07) | .292 | 0.27 (−1.39 to 1.93) | .994 | 6.04 (2.05 to 10.02) | .132 |
| Caudal middle frontal | 1.35 (−3.83 to 6.53) | .941 | −0.01 (−1.71 to 1.70) | .994 | 1.34 (−3.60 to 6.29) | .870 |
| Cuneus | 1.08 (−3.72 to 5.88) | .941 | 0.63 (−1.55 to 2.80) | .994 | 1.70 (−2.58 to 5.99) | .870 |
| Entorhinal cortex | 4.28 (−1.64 to 10.21) | .941 | 1.33 (−1.66 to 4.32) | .941 | 5.61 (0.51 to 10.71) | .218 |
| Frontal pole | 0.61 (−3.72 to 4.93) | .941 | 0.71 (−1.60 to 3.01) | .994 | 1.31 (−2.48 to 5.10) | .870 |
| Fusiform gyrus | −1.32 (−6.92 to 4.29) | .941 | 2.29 (0.46 to 4.13) | .380 | 0.98 (−4.42 to 6.38) | .870 |
| Inferior parietal | 0.17 (−5.15 to 5.48) | .974 | 0.90 (−1.21 to 3.00) | .941 | 1.06 (−3.95 to 6.07) | .870 |
| Inferior temporal | −2.72 (−7.48 to 2.04) | .941 | 1.61 (−0.86 to 4.08) | .846 | −1.11 (−5.30 to 3.08) | .870 |
| Insula | 2.96 (−1.88 to 7.81) | .941 | −0.55 (−2.46 to 1.35) | .994 | 2.41 (−2.16 to 6.98) | .870 |
| Isthmus cingulate | −5.94 (−10.94 to −0.94) | .292 | 1.78 (0.20 to 3.36) | .380 | −4.16 (−8.96 to 0.65) | .435 |
| Lateral occipital | 2.42 (−2.38 to 7.21) | .941 | 0.71 (−0.92 to 2.33) | .941 | 3.12 (−1.43 to 7.68) | .623 |
| Lateral orbitofrontal | 2.26 (−1.98 to 6.51) | .941 | −0.47 (−2.42 to 1.47) | .994 | 1.79 (−2.12 to 5.70) | .870 |
| Lingual gyrus | −0.01 (−5.83 to 5.80) | .997 | 0.48 (−1.28 to 2.23) | .994 | 0.46 (−5.07 to 6.00) | .890 |
| Medial orbitofrontal | 1.86 (−2.75 to 6.47) | .941 | −1.33 (−3.58 to 0.93) | .886 | 0.53 (−3.54 to 4.60) | .870 |
| Middle temporal | 0.98 (−3.76 to 5.72) | .941 | 1.23 (−0.90 to 3.36) | .886 | 2.21 (−2.11 to 6.53) | .870 |
| Paracentral lobule | −1.69 (−6.84 to 3.45) | .941 | 1.21 (−0.65 to 3.07) | .846 | −0.48 (−5.41 to 4.44) | .889 |
| Parahippocampal cortex | 4.69 (−1.16 to 10.54) | .941 | −0.08 (−2.24 to 2.09) | .994 | 4.61 (−0.78 to 10.01) | .435 |
| Pars opercularis | −1.20 (−6.45 to 4.05) | .941 | −0.19 (−2.30 to 1.93) | .994 | −1.39 (−6.24 to 3.46) | .870 |
| Pars orbitalis | 1.21 (−2.40 to 4.82) | .941 | −1.96 (−4.34 to 0.43) | .846 | −0.74 (−3.47 to 1.98) | .870 |
| Pars triangularis | 0.35 (−4.63 to 5.33) | .958 | −0.33 (−2.50 to 1.84) | .994 | 0.02 (−4.55 to 4.59) | .992 |
| Pericalcarine cortex | 0.90 (−4.38 to 6.18) | .941 | −0.08 (−1.89 to 1.73) | .994 | 0.82 (−4.08 to 5.72) | .870 |
| Postcentral gyrus | −1.07 (−6.94 to 4.80) | .941 | 0.01 (−1.77 to 1.79) | .994 | −1.05 (−6.83 to 4.72) | .870 |
| Posterior cingulate | −0.63 (−4.56 to 3.31) | .941 | 0.07 (−2.09 to 2.23) | .994 | −0.56 (−3.99 to 2.88) | .870 |
| Precentral gyrus | −1.34 (−7.08 to 4.39) | .941 | 0.26 (−1.44 to 1.95) | .994 | −1.09 (−6.67 to 4.50) | .870 |
| Precuneus | −0.30 (−5.11 to 4.50) | .958 | 0.96 (−1.23 to 3.15) | .941 | 0.65 (−3.75 to 5.05) | .870 |
| Rostral anterior cingulate | 5.41 (0.83 to 9.99) | .292 | −0.43 (−2.78 to 1.91) | .994 | 4.97 (1.03 to 8.92) | .169 |
| Rostral middle frontal | −0.30 (−5.69 to 5.09) | .958 | −0.30 (−2.59 to 2.00) | .994 | −0.60 (−5.42 to 4.22) | .870 |
| Superior frontal | 0.46 (−4.66 to 5.58) | .958 | 0.21 (−1.63 to 2.06) | .994 | 0.67 (−4.21 to 5.54) | .870 |
| Superior parietal | 0.65 (−4.60 to 5.91) | .941 | 0.33 (−1.56 to 2.23) | .994 | 0.98 (−4.06 to 6.03) | .870 |

| **Region** | **Cohort difference B (95% CI)** | FDR p | **PREVENT-AD B (95% CI)** | FDR p | **UCLouvain B (95% CI)** | FDR p |
|---|---|---|---|---|---|---|
| Superior temporal | −1.96 (−6.71 to 2.80) | .941 | 0.29 (−1.75 to 2.33) | .994 | −1.67 (−6.32 to 2.99) | .870 |
| Supramarginal gyrus | 1.22 (−3.95 to 6.40) | .941 | 0.41 (−1.45 to 2.26) | .994 | 1.63 (−3.41 to 6.67) | .870 |
| Temporal pole | 3.17 (−1.93 to 8.27) | .941 | 2.33 (−0.42 to 5.07) | .846 | 5.50 (1.03 to 9.96) | .169 |
| Transverse temporal | −1.50 (−6.85 to 3.85) | .941 | 0.73 (−0.93 to 2.39) | .941 | −0.77 (−5.88 to 4.34) | .870 |
| Thalamus | 3.68 (−3.05 to 10.41) | .941 | −0.09 (−2.57 to 2.38) | .994 | 3.59 (−3.28 to 10.46) | .870 |
| Caudate | 0.78 (−5.48 to 7.05) | .941 | 1.24 (−0.99 to 3.46) | .886 | 2.02 (−4.26 to 8.30) | .870 |
| Putamen | 3.07 (−1.42 to 7.55) | .941 | 1.72 (−0.85 to 4.30) | .846 | 4.79 (0.61 to 8.97) | .209 |
| Pallidum | −1.67 (−6.34 to 2.99) | .941 | −0.03 (−2.62 to 2.56) | .994 | −1.70 (−6.46 to 3.06) | .870 |
| Hippocampus | 3.48 (−1.60 to 8.55) | .941 | 0.85 (−2.29 to 3.98) | .994 | 4.33 (−0.02 to 8.67) | .307 |
| Amygdala | 1.88 (−4.19 to 7.94) | .941 | 2.37 (−0.91 to 5.65) | .846 | 4.25 (−1.89 to 10.38) | .623 |
| Accumbens area | 3.98 (−0.88 to 8.85) | .941 | 2.21 (0.37 to 4.05) | .380 | 6.19 (1.59 to 10.79) | .169 |
| Ventral diencephalon | 1.72 (−3.56 to 6.99) | .941 | 1.98 (−0.79 to 4.76) | .846 | 3.70 (−1.54 to 8.94) | .623 |

**Note.** B is the adjusted percentage-point change in odor identification score per 1 within-cohort SD increase in the biomarker. Tau-PET models included global cortical tau burden and its interactions with cohort and amyloid status; subcortical-volume models additionally included eTIV. FDR p-values are Benjamini–Hochberg corrected across all 42 regions separately by modality, group, and hypothesis. Cohort difference is UCLouvain minus PREVENT-AD.

#### Supplementary Table 3. Whole-brain amyloid-specific associations and biomarker-by-amyloid interactions

| Region | CN A−<br>B (95% CI) | FDR p | CN A+<br>B (95% CI) | FDR p | Amyloid interaction<br>B (95% CI) | FDR p |
|---|---|---|---|---|---|---|
| Tau PET | | | | | | |
| Banks of superior temporal sulcus | 0.48 (−3.20 to 4.15) | .881 | −0.86 (−5.22 to 3.50) | .998 | −1.34 (−6.87 to 4.19) | .887 |
| Caudal anterior cingulate | −0.32 (−3.69 to 3.05) | .896 | 1.53 (−3.42 to 6.49) | .950 | 1.85 (−4.16 to 7.86) | .881 |
| Caudal middle frontal | 4.10 (−1.35 to 9.55) | .487 | −0.08 (−6.92 to 6.77) | .998 | −4.18 (−13.24 to 4.88) | .779 |
| Cuneus | 2.59 (−1.20 to 6.38) | .502 | 1.77 (−1.56 to 5.10) | .820 | −0.82 (−6.10 to 4.45) | .923 |
| Entorhinal cortex | 1.96 (−0.99 to 4.92) | .504 | 0.13 (−3.15 to 3.41) | .998 | −1.84 (−6.03 to 2.36) | .779 |
| Frontal pole | −0.56 (−3.39 to 2.26) | .858 | 1.83 (−1.90 to 5.57) | .820 | 2.40 (−2.27 to 7.07) | .779 |
| Fusiform gyrus | −1.17 (−4.91 to 2.58) | .811 | −2.52 (−6.39 to 1.35) | .820 | −1.36 (−6.67 to 3.96) | .887 |
| Inferior parietal | −2.84 (−6.84 to 1.16) | .489 | −4.83 (−13.04 to 3.39) | .820 | −1.99 (−11.12 to 7.14) | .906 |
| Inferior temporal | −0.84 (−4.08 to 2.40) | .839 | −3.25 (−8.27 to 1.78) | .820 | −2.41 (−8.38 to 3.56) | .784 |
| Insula | 3.98 (1.65 to 6.30) | **.037** | 1.25 (−5.84 to 8.33) | .998 | −2.73 (−10.27 to 4.81) | .829 |
| Isthmus cingulate | 1.34 (−2.05 to 4.73) | .743 | 0.85 (−2.47 to 4.16) | .994 | −0.49 (−5.33 to 4.35) | .923 |
| Lateral occipital | −0.71 (−4.63 to 3.21) | .866 | −0.23 (−3.02 to 2.56) | .998 | 0.48 (−4.47 to 5.43) | .923 |
| Lateral orbitofrontal | 1.55 (−1.02 to 4.12) | .524 | −4.02 (−10.81 to 2.77) | .820 | −5.57 (−12.89 to 1.75) | .779 |
| Lingual gyrus | 1.11 (−1.85 to 4.06) | .743 | 0.93 (−3.47 to 5.32) | .998 | −0.18 (−5.62 to 5.26) | .948 |
| Medial orbitofrontal | 1.28 (−1.97 to 4.54) | .743 | 1.76 (−3.55 to 7.07) | .950 | 0.48 (−5.86 to 6.82) | .923 |
| Middle temporal | 2.00 (−3.25 to 7.25) | .743 | −1.46 (−6.18 to 3.25) | .950 | −3.47 (−10.53 to 3.60) | .779 |
| Paracentral lobule | 1.25 (−1.47 to 3.98) | .738 | 2.83 (−2.28 to 7.94) | .820 | 1.57 (−4.24 to 7.39) | .887 |
| Parahippocampal cortex | 2.02 (−1.29 to 5.34) | .524 | −0.13 (−4.60 to 4.35) | .998 | −2.15 (−7.51 to 3.20) | .784 |
| Pars opercularis | 0.45 (−2.88 to 3.78) | .881 | 3.96 (−2.74 to 10.67) | .820 | 3.51 (−3.74 to 10.76) | .779 |
| Pars orbitalis | −0.45 (−3.21 to 2.30) | .869 | −0.01 (−5.33 to 5.31) | .998 | 0.45 (−5.50 to 6.39) | .923 |
| Pars triangularis | 0.64 (−1.83 to 3.11) | .839 | 3.89 (−0.69 to 8.47) | .820 | 3.25 (−1.93 to 8.44) | .779 |
| Pericalcarine cortex | 0.33 (−2.52 to 3.18) | .881 | 3.41 (−0.11 to 6.93) | .820 | 3.08 (−1.58 to 7.74) | .779 |
| Postcentral gyrus | −0.90 (−4.47 to 2.67) | .839 | 3.02 (−2.50 to 8.55) | .820 | 3.93 (−2.76 to 10.61) | .779 |
| Posterior cingulate | 1.34 (−2.25 to 4.93) | .743 | −0.36 (−5.54 to 4.81) | .998 | −1.71 (−7.98 to 4.57) | .887 |
| Precentral gyrus | 3.34 (−0.06 to 6.74) | .264 | 4.14 (−1.70 to 9.97) | .820 | 0.80 (−6.03 to 7.63) | .923 |
| Precuneus | 0.15 (−6.23 to 6.53) | .987 | 0.85 (−4.08 to 5.78) | .998 | 0.70 (−7.60 to 9.00) | .923 |
| Rostral anterior cingulate | −0.50 (−2.82 to 1.81) | .858 | 2.72 (−1.82 to 7.26) | .820 | 3.22 (−1.85 to 8.29) | .779 |
| Rostral middle frontal | −1.60 (−4.25 to 1.06) | .524 | 2.39 (−4.42 to 9.19) | .950 | 3.99 (−3.35 to 11.32) | .779 |
| Superior frontal | −1.95 (−6.23 to 2.32) | .738 | 1.00 (−6.16 to 8.17) | .998 | 2.96 (−5.53 to 11.45) | .829 |
| Superior parietal | −1.08 (−4.08 to 1.92) | .743 | −1.71 (−7.76 to 4.34) | .970 | −0.63 (−7.28 to 6.02) | .923 |
| Superior temporal | 0.68 (−2.71 to 4.07) | .858 | −0.82 (−9.00 to 7.36) | .998 | −1.50 (−9.55 to 6.55) | .923 |
| Supramarginal gyrus | −3.51 (−7.50 to 0.47) | .320 | −2.97 (−10.81 to 4.88) | .950 | 0.54 (−8.02 to 9.11) | .923 |
| Temporal pole | 0.01 (−2.56 to 2.57) | .997 | −6.93 (−11.58 to −2.28) | .153 | −6.93 (−12.17 to −1.70) | .273 |
| Transverse temporal | 1.58 (−0.63 to 3.79) | .489 | 3.62 (−0.59 to 7.82) | .820 | 2.04 (−2.55 to 6.63) | .779 |
| Thalamus | 2.05 (0.21 to 3.89) | .188 | −0.99 (−6.81 to 4.83) | .998 | −3.04 (−9.04 to 2.97) | .779 |
| Caudate | 2.58 (0.76 to 4.40) | .120 | 0.40 (−3.43 to 4.23) | .998 | −2.18 (−6.49 to 2.13) | .779 |
| Putamen | 2.25 (−0.08 to 4.58) | .264 | −2.65 (−7.94 to 2.63) | .820 | −4.90 (−10.62 to 0.81) | .775 |
| Pallidum | 2.58 (0.36 to 4.81) | .188 | −4.20 (−9.20 to 0.79) | .820 | −6.78 (−12.13 to −1.44) | .273 |
| Hippocampus | 2.44 (0.28 to 4.60) | .188 | −0.28 (−4.96 to 4.41) | .998 | −2.72 (−7.89 to 2.46) | .779 |
| Amygdala | 3.14 (0.28 to 5.99) | .188 | −1.38 (−4.41 to 1.65) | .820 | −4.52 (−8.48 to −0.56) | .358 |
| Accumbens area | 2.52 (0.54 to 4.50) | .180 | 0.02 (−4.78 to 4.82) | .998 | −2.50 (−7.41 to 2.41) | .779 |
| Ventral diencephalon | 2.52 (−0.14 to 5.18) | .264 | −2.07 (−6.63 to 2.48) | .820 | −4.60 (−9.84 to 0.64) | .775 |
| Morphometry | | | | | | |
| Banks of superior temporal sulcus | 0.21 (−1.85 to 2.27) | .914 | 0.94 (−2.70 to 4.58) | .903 | 0.73 (−3.40 to 4.87) | .872 |
| Caudal anterior cingulate | 0.49 (−1.04 to 2.03) | .914 | 2.58 (−0.96 to 6.12) | .534 | 2.08 (−1.73 to 5.90) | .813 |
| Caudal middle frontal | 0.51 (−1.40 to 2.42) | .914 | −0.41 (−3.59 to 2.76) | .943 | −0.92 (−4.61 to 2.77) | .831 |
| Cuneus | 0.21 (−1.92 to 2.34) | .914 | 1.86 (−1.98 to 5.71) | .720 | 1.65 (−2.74 to 6.04) | .813 |
| Entorhinal cortex | −0.40 (−2.28 to 1.47) | .914 | 4.79 (0.43 to 9.15) | .329 | 5.19 (0.38 to 10.00) | .453 |
| Frontal pole | 0.78 (−1.81 to 3.37) | .914 | 0.83 (−2.50 to 4.16) | .903 | 0.05 (−4.11 to 4.20) | .997 |
| Fusiform gyrus | 1.09 (−0.57 to 2.74) | .914 | 3.59 (−0.10 to 7.27) | .402 | 2.50 (−1.46 to 6.46) | .813 |
| Inferior parietal | 0.21 (−1.94 to 2.36) | .914 | 2.10 (−1.48 to 5.69) | .720 | 1.89 (−2.12 to 5.91) | .813 |
| Inferior temporal | 1.85 (−0.75 to 4.44) | .914 | 0.33 (−2.74 to 3.41) | .943 | −1.51 (−5.45 to 2.42) | .813 |
| Insula | −0.39 (−2.45 to 1.66) | .914 | 0.61 (−2.51 to 3.73) | .919 | 1.00 (−2.50 to 4.51) | .813 |
| Isthmus cingulate | 0.85 (−0.83 to 2.53) | .914 | 0.99 (−2.57 to 4.55) | .903 | 0.14 (−3.75 to 4.03) | .997 |
| Lateral occipital | 0.15 (−1.00 to 1.30) | .914 | 2.91 (−1.21 to 7.03) | .535 | 2.76 (−1.47 to 6.99) | .813 |
| Lateral orbitofrontal | −0.55 (−2.42 to 1.32) | .914 | 0.82 (−2.91 to 4.55) | .917 | 1.37 (−2.61 to 5.35) | .813 |
| Lingual gyrus | −0.37 (−1.85 to 1.11) | .914 | 2.04 (−2.01 to 6.09) | .720 | 2.41 (−1.92 to 6.74) | .813 |
| Medial orbitofrontal | −0.70 (−2.68 to 1.27) | .914 | −1.57 (−5.55 to 2.41) | .799 | −0.87 (−5.20 to 3.47) | .857 |
| Middle temporal | 0.79 (−0.89 to 2.46) | .914 | 2.23 (−2.07 to 6.53) | .720 | 1.44 (−3.04 to 5.93) | .813 |
| Paracentral lobule | 0.52 (−1.19 to 2.24) | .914 | 2.04 (−2.19 to 6.27) | .720 | 1.52 (−2.94 to 5.97) | .813 |
| Parahippocampal cortex | −1.29 (−2.99 to 0.41) | .914 | 4.86 (−0.34 to 10.07) | .402 | 6.15 (0.56 to 11.74) | .453 |
| Pars opercularis | 0.40 (−1.55 to 2.36) | .914 | −1.79 (−5.76 to 2.19) | .755 | −2.19 (−6.61 to 2.23) | .813 |
| Pars orbitalis | −0.98 (−2.93 to 0.96) | .914 | −3.57 (−8.22 to 1.08) | .503 | −2.59 (−7.50 to 2.33) | .813 |
| Pars triangularis | 0.30 (−1.93 to 2.54) | .914 | −1.83 (−5.35 to 1.70) | .720 | −2.13 (−6.20 to 1.94) | .813 |
| Pericalcarine cortex | −0.26 (−2.40 to 1.87) | .914 | 0.59 (−2.20 to 3.38) | .917 | 0.86 (−2.67 to 4.38) | .831 |
| Postcentral gyrus | −0.32 (−1.99 to 1.35) | .914 | 0.20 (−4.11 to 4.52) | .990 | 0.52 (−4.03 to 5.07) | .933 |
| Posterior cingulate | 0.27 (−1.93 to 2.46) | .914 | −1.18 (−4.79 to 2.43) | .842 | −1.44 (−5.55 to 2.66) | .813 |
| Precentral gyrus | 0.10 (−1.65 to 1.85) | .944 | −0.08 (−3.81 to 3.65) | .990 | −0.18 (−4.23 to 3.87) | .997 |
| Precuneus | 0.42 (−1.25 to 2.08) | .914 | 1.67 (−2.54 to 5.87) | .799 | 1.25 (−3.20 to 5.70) | .813 |
| Rostral anterior cingulate | 0.82 (−1.72 to 3.35) | .914 | −0.16 (−3.94 to 3.62) | .990 | −0.98 (−5.56 to 3.61) | .857 |
| Rostral middle frontal | 1.28 (−1.00 to 3.56) | .914 | −2.85 (−5.81 to 0.11) | .402 | −4.13 (−7.83 to −0.43) | .453 |
| Superior frontal | 0.62 (−1.45 to 2.70) | .914 | −0.51 (−3.54 to 2.51) | .927 | −1.13 (−4.69 to 2.43) | .813 |
| Superior parietal | −0.08 (−1.75 to 1.58) | .944 | 1.47 (−2.72 to 5.66) | .823 | 1.55 (−2.89 to 6.00) | .813 |

| Region | CN A−<br>B (95% CI) | FDR p | CN A+<br>B (95% CI) | FDR p | Amyloid interaction<br>B (95% CI) | FDR p |
|---|---|---|---|---|---|---|
| Superior temporal | −0.01 (−1.80 to 1.77) | .988 | −0.02 (−4.01 to 3.97) | .991 | −0.01 (−4.04 to 4.02) | .997 |
| Supramarginal gyrus | 0.19 (−1.62 to 2.00) | .914 | 1.36 (−2.49 to 5.21) | .823 | 1.17 (−2.92 to 5.26) | .813 |
| Temporal pole | 1.17 (−1.14 to 3.48) | .914 | 5.28 (0.55 to 10.01) | .329 | 4.11 (−1.10 to 9.32) | .769 |
| Transverse temporal | 0.49 (−0.98 to 1.95) | .914 | 0.50 (−3.72 to 4.72) | .943 | 0.02 (−4.48 to 4.51) | .997 |
| Thalamus | 0.64 (−1.77 to 3.04) | .914 | −0.11 (−4.80 to 4.58) | .990 | −0.75 (−5.59 to 4.10) | .889 |
| Caudate | 0.78 (−1.72 to 3.28) | .914 | 2.10 (−1.78 to 5.97) | .720 | 1.31 (−3.05 to 5.68) | .813 |
| Putamen | 1.72 (−0.73 to 4.18) | .914 | 4.21 (−0.90 to 9.32) | .480 | 2.49 (−2.76 to 7.74) | .813 |
| Pallidum | −0.57 (−3.16 to 2.02) | .914 | 0.69 (−3.59 to 4.97) | .927 | 1.26 (−2.87 to 5.39) | .813 |
| Hippocampus | −0.31 (−2.67 to 2.05) | .914 | 4.23 (−1.03 to 9.50) | .480 | 4.55 (−0.93 to 10.02) | .769 |
| Amygdala | 0.54 (−1.91 to 2.99) | .914 | 5.64 (0.55 to 10.73) | .329 | 5.10 (0.16 to 10.04) | .453 |
| Accumbens area | 1.91 (0.29 to 3.54) | .884 | 5.66 (0.94 to 10.38) | .329 | 3.75 (−1.09 to 8.58) | .769 |
| Ventral diencephalon | 1.67 (−0.79 to 4.12) | .914 | 5.38 (−0.88 to 11.64) | .480 | 3.71 (−2.03 to 9.45) | .813 |

**Note.** B is the adjusted percentage-point change in odor identification score per 1 within-cohort SD increase in the biomarker. Tau-PET models included global cortical tau burden and its interactions with cohort and amyloid status; subcortical-volume models additionally included eTIV. FDR p values are Benjamini–Hochberg corrected across all 42 regions separately by modality, group, and hypothesis. Amyloid interaction is CN A+ minus CN A−.

#### Supplementary Table 4. Whole-brain biomarker-by-cohort-by-amyloid interactions

| Region | Cohort × amyloid interaction B (95% CI) | p | FDR p | n |
|---|---|---|---|---|
| Tau PET | | | | |
| Banks of superior temporal sulcus | −41.98 (−82.86 to −1.10) | **.044** | .232 | 245 |
| Caudal anterior cingulate | −3.75 (−17.85 to 10.35) | .601 | .720 | 245 |
| Caudal middle frontal | −17.97 (−35.31 to −0.63) | **.042** | .232 | 245 |
| Cuneus | −6.71 (−20.85 to 7.43) | .351 | .546 | 245 |
| Entorhinal cortex | −10.67 (−23.22 to 1.89) | .096 | .334 | 245 |
| Frontal pole | 1.77 (−11.86 to 15.39) | .798 | .860 | 245 |
| Fusiform gyrus | −8.29 (−21.50 to 4.92) | .217 | .435 | 245 |
| Inferior parietal | 2.78 (−42.63 to 48.20) | .904 | .926 | 245 |
| Inferior temporal | 2.69 (−23.85 to 29.22) | .842 | .884 | 245 |
| Insula | −16.77 (−31.14 to −2.39) | **.022** | .157 | 245 |
| Isthmus cingulate | −18.80 (−44.82 to 7.23) | .156 | .403 | 245 |
| Lateral occipital | −0.52 (−17.78 to 16.74) | .953 | .953 | 245 |
| Lateral orbitofrontal | −2.61 (−15.53 to 10.30) | .691 | .764 | 245 |
| Lingual gyrus | −12.84 (−21.13 to −4.55) | **.003** | .074 | 245 |
| Medial orbitofrontal | −9.30 (−19.84 to 1.23) | .083 | .328 | 245 |
| Middle temporal | −13.78 (−46.56 to 18.99) | .408 | .554 | 245 |
| Paracentral lobule | −2.74 (−13.50 to 8.03) | .617 | .720 | 245 |
| Parahippocampal cortex | −6.85 (−24.12 to 10.42) | .435 | .571 | 245 |
| Pars opercularis | −21.06 (−35.82 to −6.30) | **.005** | .075 | 245 |
| Pars orbitalis | 5.39 (−7.44 to 18.21) | .409 | .554 | 245 |
| Pars triangularis | 6.71 (−2.98 to 16.39) | .174 | .405 | 245 |
| Pericalcarine cortex | −4.72 (−15.70 to 6.27) | .398 | .554 | 245 |
| Postcentral gyrus | 4.18 (−7.24 to 15.60) | .471 | .600 | 245 |
| Posterior cingulate | −9.61 (−26.02 to 6.80) | .250 | .477 | 245 |
| Precentral gyrus | 3.09 (−7.17 to 13.35) | .554 | .684 | 245 |
| Precuneus | −36.91 (−67.51 to −6.30) | **.018** | .154 | 245 |
| Rostral anterior cingulate | −5.56 (−15.79 to 4.67) | .285 | .500 | 245 |
| Rostral middle frontal | 5.83 (−4.89 to 16.55) | .285 | .500 | 245 |
| Superior frontal | −7.14 (−15.30 to 1.01) | .086 | .328 | 245 |
| Superior parietal | −11.92 (−30.06 to 6.22) | .197 | .430 | 245 |
| Superior temporal | −11.56 (−27.85 to 4.73) | .163 | .403 | 245 |
| Supramarginal gyrus | −15.22 (−35.74 to 5.30) | .145 | .403 | 245 |
| Temporal pole | −8.46 (−19.44 to 2.53) | .131 | .392 | 245 |
| Transverse temporal | −6.82 (−17.40 to 3.75) | .205 | .430 | 245 |
| Thalamus | 2.30 (−8.41 to 13.01) | .672 | .763 | 245 |
| Caudate | −11.04 (−22.11 to 0.03) | .051 | .236 | 245 |
| Putamen | 5.93 (−8.12 to 19.98) | .407 | .554 | 245 |
| Pallidum | −8.42 (−25.66 to 8.82) | .337 | .544 | 245 |
| Hippocampus | −19.73 (−32.92 to −6.54) | **.004** | .074 | 245 |
| Amygdala | −24.19 (−54.49 to 6.11) | .117 | .378 | 245 |
| Accumbens area | −18.50 (−32.48 to −4.52) | **.010** | .102 | 245 |
| Ventral diencephalon | −9.34 (−27.11 to 8.43) | .302 | .507 | 245 |
| Morphometry | | | | |
| Banks of superior temporal sulcus | 2.82 (−7.33 to 12.98) | .585 | .912 | 248 |
| Caudal anterior cingulate | 4.80 (−3.41 to 13.01) | .251 | .658 | 248 |
| Caudal middle frontal | 9.55 (−1.01 to 20.11) | .076 | .406 | 248 |
| Cuneus | −0.56 (−10.34 to 9.22) | .910 | .984 | 248 |
| Entorhinal cortex | −3.45 (−15.47 to 8.57) | .572 | .912 | 248 |
| Frontal pole | 0.15 (−11.98 to 12.29) | .980 | .984 | 248 |
| Fusiform gyrus | 1.70 (−8.98 to 12.38) | .754 | .984 | 248 |
| Inferior parietal | 7.07 (−2.31 to 16.46) | .139 | .534 | 248 |
| Inferior temporal | 3.18 (−7.47 to 13.83) | .557 | .912 | 248 |
| Insula | −1.35 (−14.77 to 12.06) | .842 | .984 | 248 |
| Isthmus cingulate | −7.29 (−17.11 to 2.53) | .145 | .534 | 248 |
| Lateral occipital | 9.09 (−0.53 to 18.70) | .064 | .406 | 248 |
| Lateral orbitofrontal | 7.18 (−6.75 to 21.12) | .311 | .768 | 248 |
| Lingual gyrus | 0.40 (−11.13 to 11.92) | .946 | .984 | 248 |
| Medial orbitofrontal | 2.10 (−8.21 to 12.41) | .689 | .984 | 248 |
| Middle temporal | 9.91 (−0.81 to 20.63) | .070 | .406 | 248 |
| Paracentral lobule | −0.17 (−14.02 to 13.69) | .981 | .984 | 248 |
| Parahippocampal cortex | 2.90 (−7.53 to 13.32) | .585 | .912 | 248 |
| Pars opercularis | 4.59 (−8.83 to 18.01) | .501 | .912 | 248 |
| Pars orbitalis | 4.69 (−8.07 to 17.44) | .470 | .912 | 248 |
| Pars triangularis | 16.99 (3.15 to 30.84) | **.016** | .406 | 248 |
| Pericalcarine cortex | −1.62 (−11.29 to 8.05) | .741 | .984 | 248 |
| Postcentral gyrus | 1.64 (−12.03 to 15.32) | .813 | .984 | 248 |
| Posterior cingulate | −17.45 (−33.38 to −1.51) | **.032** | .406 | 248 |
| Precentral gyrus | −0.14 (−13.45 to 13.17) | .984 | .984 | 248 |
| Precuneus | 0.81 (−9.60 to 11.22) | .879 | .984 | 248 |
| Rostral anterior cingulate | 7.97 (−1.17 to 17.10) | .087 | .406 | 248 |
| Rostral middle frontal | 5.64 (−8.08 to 19.35) | .419 | .912 | 248 |

| Region | Cohort × amyloid interaction<br>B (95% CI) | p | FDR p | n |
|---|---|---|---|---|
| Superior frontal | 5.99 (−7.67 to 19.65) | .389 | .907 | 248 |
| Superior parietal | 7.75 (−3.53 to 19.03) | .177 | .544 | 248 |
| Superior temporal | −1.75 (−15.68 to 12.17) | .804 | .984 | 248 |
| Supramarginal gyrus | 7.40 (−2.76 to 17.56) | .152 | .534 | 248 |
| Temporal pole | 5.87 (−3.47 to 15.20) | .217 | .608 | 248 |
| Transverse temporal | −3.41 (−15.64 to 8.83) | .584 | .912 | 248 |
| Thalamus | −11.77 (−24.50 to 0.97) | .070 | .406 | 239 |
| Caudate | 11.37 (−1.55 to 24.29) | .084 | .406 | 248 |
| Putamen | 3.23 (−8.44 to 14.89) | .586 | .912 | 248 |
| Pallidum | −1.78 (−11.80 to 8.24) | .727 | .984 | 248 |
| Hippocampus | −9.69 (−23.94 to 4.56) | .181 | .544 | 248 |
| Amygdala | −0.85 (−13.99 to 12.28) | .898 | .984 | 248 |
| Accumbens area | −0.73 (−13.61 to 12.16) | .911 | .984 | 248 |
| Ventral diencephalon | −13.07 (−25.60 to −0.54) | **.041** | .406 | 248 |

**Note.** The coefficient is the biomarker-by-cohort-by-amyloid interaction. These models are exploratory. Tau-PET models included global cortical tau burden and its interactions with cohort and amyloid status; subcortical-volume models additionally included eTIV. FDR p-values are Benjamini–Hochberg corrected across all 42 regions separately by modality.

**Supplementary Table 5. Exploratory visual tau-positivity analysis in UCLouvain**

| Visual tau status | n | Odor identification, mean (SD), % | Adjusted difference B (95% CI) | p |
|---|---|---|---|---|
| Tau negative | 20 | 73.4 (19.1) | Reference | — |
| Tau positive | 11 | 56.2 (19.8) | −13.90 (−30.56 to 2.76) | .098 |

Note. Visual tau positivity was defined as a Braak-like visual stage greater than 0. The adjusted difference compares tau-positive with tau-negative participants in a linear model adjusted for amyloid status, age, sex, education, and APOE ε4-carrier status. The confidence interval uses an HC1 robust standard error. This analysis was exploratory and was not included in an FDR family.

#### Supplementary Table 6. Tau-PET sensitivity analyses by olfaction-to-PET interval

| Window | Region | Cohort difference B (95% CI) | FDR p | PREVENT-AD B (95% CI) | FDR p | UCLouvain B (95% CI) | FDR p | n (P/U) |
|---|---|---|---|---|---|---|---|---|
| Full sample | Amygdala | −7.41 (−13.38 to −1.44) | **.030** | 0.82 (−1.42 to 3.06) | .471 | −6.59 (−12.08 to −1.10) | **.038** | 215/30 |
| Full sample | Hippocampus | −5.95 (−15.76 to 3.86) | .234 | 1.42 (−1.42 to 4.27) | .434 | −4.52 (−13.90 to 4.85) | .343 | 215/30 |
| Full sample | Entorhinal cortex | −11.55 (−22.15 to −0.96) | **.044** | 1.27 (−1.08 to 3.63) | .434 | −10.28 (−20.54 to −0.02) | .066 | 215/30 |
| Full sample | Parahippocampal cortex | −14.49 (−22.74 to −6.24) | **.003** | 1.68 (−0.96 to 4.33) | .434 | −12.81 (−20.42 to −5.20) | **.004** | 215/30 |
| Within 24 months | Amygdala | −6.33 (−12.96 to 0.30) | .082 | −0.14 (−3.80 to 3.52) | .939 | −6.47 (−12.11 to −0.83) | **.033** | 119/23 |
| Within 24 months | Hippocampus | −5.86 (−14.12 to 2.39) | .162 | −1.01 (−4.75 to 2.74) | .939 | −6.87 (−14.23 to 0.48) | .067 | 119/23 |
| Within 24 months | Entorhinal cortex | −11.49 (−21.31 to −1.67) | **.044** | −0.86 (−3.16 to 1.44) | .939 | −12.35 (−21.65 to −3.04) | **.019** | 119/23 |
| Within 24 months | Parahippocampal cortex | −15.12 (−23.65 to −6.58) | **.003** | 0.21 (−3.19 to 3.60) | .939 | −14.91 (−22.25 to −7.56) | **<.001** | 119/23 |
| Within 12 months | Amygdala | −5.49 (−12.16 to 1.18) | .141 | −1.65 (−5.70 to 2.40) | .570 | −7.14 (−12.40 to −1.88) | **.015** | 90/17 |
| Within 12 months | Hippocampus | −6.23 (−14.62 to 2.16) | .144 | −1.77 (−6.17 to 2.64) | .570 | −8.00 (−15.22 to −0.77) | **.030** | 90/17 |
| Within 12 months | Entorhinal cortex | −10.31 (−20.89 to 0.28) | .112 | −2.41 (−4.95 to 0.12) | .247 | −12.72 (−22.46 to −2.98) | **.015** | 90/17 |
| Within 12 months | Parahippocampal cortex | −13.04 (−22.68 to −3.40) | **.034** | −0.86 (−4.99 to 3.27) | .680 | −13.90 (−22.37 to −5.44) | **.006** | 90/17 |

Note. P/U denotes PREVENT-AD/UCLouvain sample sizes. The full-sample models reproduce the primary analyses. Interval-restricted analyses included participants whose olfactory and tau-PET assessments were within the specified absolute interval. Models used the same APOE ε4-adjusted covariate set as the primary tau-PET analyses. Cohort difference is UCLouvain minus PREVENT-AD. FDR p values are corrected across the four prespecified MTL regions separately by cohort, window, and hypothesis.

**Supplementary Table 7. Sensitivity analysis: adjusted MTL biomarker-by-cohort interactions and follow-up cohort-specific associations**

| Modality | Region | Cohort difference, B (95% CI) | FDR p | PREVENT-AD, B (95% CI) | FDR p | UCLouvain, B (95% CI) | FDR p |
|---|---|---|---|---|---|---|---|
| Tau PET | Amygdala | 7.77 (−9.02 to 24.57) | .388 | 0.97 (−2.06 to 4.00) | .529 | 8.74 (−7.76 to 25.25) | .297 |
| | Hippocampus | 5.61 (−7.16 to 18.37) | .388 | 1.91 (0.06 to 3.75) | .171 | 7.51 (−5.20 to 20.22) | .297 |
| | Entorhinal cortex | −11.22 (−22.12 to −0.31) | .088 | 1.04 (−1.67 to 3.75) | .529 | −10.18 (−20.70 to 0.33) | .115 |
| | Parahippocampal cortex | −15.25 (−23.82 to −6.68) | **.002** | 1.89 (−1.04 to 4.81) | .412 | −13.36 (−21.20 to −5.52) | **.004** |
| Morphometry | Amygdala | 1.87 (−4.23 to 7.97) | .547 | 2.38 (−1.00 to 5.76) | .668 | 4.25 (−1.89 to 10.38) | .174 |
| | Hippocampus | 4.69 (−0.12 to 9.49) | .112 | −0.60 (−2.84 to 1.63) | .795 | 4.08 (−0.25 to 8.41) | .126 |
| | Entorhinal cortex | 6.25 (0.69 to 11.81) | .111 | −0.61 (−2.53 to 1.31) | .795 | 5.64 (0.52 to 10.77) | .124 |
| | Parahippocampal cortex | 4.44 (−1.44 to 10.32) | .185 | 0.16 (−2.11 to 2.42) | .891 | 4.60 (−0.79 to 9.98) | .126 |

**Note.** Sensitivity analysis excluding regional imaging values beyond ±3 within-cohort SD; tau-PET models also excluded extreme global cortical tau values, and subcortical-volume models excluded extreme eTIV values. B is the adjusted percentage-point change in odor identification score per 1 within-cohort SD higher biomarker value. Cohort difference is UCLouvain minus PREVENT-AD. Tau-PET models included global cortical tau and its interactions with cohort and amyloid status. Confidence intervals use HC1 robust standard errors. FDR p-values are corrected across the four prespecified MTL regions separately by modality, cohort, and hypothesis.

#### Supplementary Table 8. Sensitivity analysis: adjusted MTL amyloid-specific associations and interaction tests

| Modality | Region | CN A−, B (95% CI) | CN A+, B (95% CI) | Amyloid interaction, B (95% CI) | FDR p | Cohort × amyloid interaction, B (95% CI) | FDR p |
|---|---|---|---|---|---|---|---|
| Tau PET | Amygdala | 3.25 (0.28 to 6.22) | −0.91 (−5.60 to 3.77) | −4.16 (−9.64 to 1.31) | .450 | −24.66 (−66.71 to 17.39) | .332 |
| | Hippocampus | 2.46 (0.30 to 4.63) | 1.41 (−1.93 to 4.76) | −1.05 (−5.03 to 2.94) | .605 | −17.30 (−38.00 to 3.40) | .202 |
| | Entorhinal cortex | 1.86 (−1.10 to 4.82) | −0.54 (−4.74 to 3.67) | −2.40 (−7.31 to 2.52) | .450 | −17.00 (−34.40 to 0.40) | .202 |
| | Parahippocampal cortex | 2.59 (−0.86 to 6.05) | −0.63 (−5.83 to 4.57) | −3.22 (−9.26 to 2.82) | .450 | −6.67 (−29.72 to 16.37) | .569 |
| Morphometry | Amygdala | 0.44 (−2.08 to 2.96) | 5.64 (0.54 to 10.73) | 5.20 (0.24 to 10.16) | .080 | −0.98 (−14.13 to 12.17) | .996 |
| | Hippocampus | −0.67 (−3.10 to 1.76) | 1.91 (−1.89 to 5.70) | 2.57 (−1.91 to 7.06) | .259 | −7.76 (−21.65 to 6.12) | .996 |
| | Entorhinal cortex | −0.38 (−2.26 to 1.51) | 2.60 (−1.70 to 6.89) | 2.98 (−1.82 to 7.77) | .259 | 0.03 (−11.92 to 11.98) | .996 |
| | Parahippocampal cortex | −1.32 (−3.03 to 0.38) | 6.44 (1.45 to 11.44) | 7.77 (2.37 to 13.16) | **.020** | 0.88 (−9.66 to 11.42) | .996 |

**Note.** Sensitivity analysis excluding imaging values beyond ±3 within-cohort SD. CN A− and CN A+ are adjusted biomarker slopes. Amyloid interaction is CN A+ minus CN A−. Cohort × amyloid interaction is the biomarker-by-cohort-by-amyloid coefficient and is exploratory. Tau-PET models included global cortical tau and its interactions with cohort and amyloid status; subcortical-volume models included eTIV. FDR p-values are corrected across the four prespecified MTL regions separately by modality and hypothesis.

### Supplementary Table 9. Sensitivity analysis: atlas-wide biomarker-by-cohort interactions and follow-up cohort-specific associations

| **Modality** | **Region** | **Cohort difference, B (95% CI)** | FDR p | **PREVENT-AD, B (95% CI)** | FDR p | **UCLouvain, B (95% CI)** | FDR p |
|---|---|---|---|---|---|---|---|
| Tau PET | Accumbens area | −8.55 (−29.58 to 12.47) | .712 | 2.11 (0.04 to 4.18) | .418 | −6.44 (−27.43 to 14.55) | .819 |
| Tau PET | Amygdala | 7.77 (−9.02 to 24.57) | .662 | 0.97 (−2.06 to 4.00) | .831 | 8.74 (−7.76 to 25.25) | .568 |
| Tau PET | Banks of superior temporal sulcus | −3.92 (−72.94 to 65.10) | .986 | 0.30 (−3.28 to 3.88) | .956 | −3.62 (−72.46 to 65.21) | .999 |
| Tau PET | Caudal anterior cingulate | 4.89 (−9.54 to 19.32) | .816 | −0.16 (−3.10 to 2.78) | .956 | 4.73 (−9.42 to 18.88) | .795 |
| Tau PET | Caudal middle frontal | 41.07 (14.09 to 68.06) | **.021** | 2.26 (−2.19 to 6.71) | .702 | 43.33 (16.58 to 70.09) | **.010** |
| Tau PET | Caudate | −0.69 (−14.70 to 13.33) | .986 | 1.60 (−0.15 to 3.36) | .418 | 0.92 (−12.95 to 14.78) | .999 |
| Tau PET | Cuneus | 0.26 (−14.52 to 15.03) | .986 | 2.71 (−0.89 to 6.31) | .487 | 2.97 (−10.92 to 16.85) | .976 |
| Tau PET | Entorhinal cortex | −11.22 (−22.12 to −0.31) | .167 | 1.04 (−1.67 to 3.75) | .779 | −10.18 (−20.70 to 0.33) | .220 |
| Tau PET | Frontal pole | 7.92 (−3.45 to 19.29) | .423 | −1.30 (−4.23 to 1.64) | .749 | 6.62 (−4.20 to 17.44) | .481 |
| Tau PET | Fusiform gyrus | −13.76 (−20.85 to −6.66) | **.004** | −0.81 (−4.04 to 2.42) | .831 | −14.56 (−20.73 to −8.40) | **<.001** |
| Tau PET | Hippocampus | 5.61 (−7.16 to 18.37) | .679 | 1.91 (0.06 to 3.75) | .418 | 7.51 (−5.20 to 20.22) | .491 |
| Tau PET | Inferior parietal | −19.36 (−54.31 to 15.59) | .580 | −4.43 (−8.99 to 0.13) | .418 | −23.79 (−58.66 to 11.08) | .421 |
| Tau PET | Inferior temporal | −41.56 (−58.64 to −24.48) | **<.001** | −3.22 (−8.13 to 1.68) | .560 | −44.78 (−61.30 to −28.27) | **<.001** |
| Tau PET | Insula | −5.40 (−39.93 to 29.14) | .986 | 2.93 (−0.14 to 6.00) | .418 | −2.47 (−36.93 to 32.00) | .999 |
| Tau PET | Isthmus cingulate | 1.13 (−34.34 to 36.61) | .986 | 0.90 (−1.52 to 3.32) | .779 | 2.03 (−33.28 to 37.35) | .999 |
| Tau PET | Lateral occipital | −18.92 (−57.77 to 19.92) | .662 | 1.97 (−0.37 to 4.31) | .456 | −16.95 (−55.66 to 21.76) | .681 |
| Tau PET | Lateral orbitofrontal | 2.72 (−25.49 to 30.93) | .986 | 0.20 (−2.67 to 3.08) | .956 | 2.92 (−25.02 to 30.86) | .999 |
| Tau PET | Lingual gyrus | −2.05 (−20.22 to 16.12) | .986 | 2.12 (−0.25 to 4.48) | .418 | 0.06 (−17.91 to 18.04) | .999 |
| Tau PET | Medial orbitofrontal | 22.34 (1.93 to 42.75) | .143 | 0.27 (−2.63 to 3.16) | .956 | 22.60 (2.38 to 42.83) | .134 |
| Tau PET | Middle temporal | −16.46 (−135.37 to 102.45) | .986 | 0.77 (−4.90 to 6.44) | .956 | −15.69 (−134.38 to 103.00) | .999 |
| Tau PET | Pallidum | 0.13 (−14.14 to 14.39) | .986 | −0.12 (−2.83 to 2.60) | .956 | 0.01 (−14.06 to 14.08) | .999 |
| Tau PET | Paracentral lobule | 9.51 (−0.85 to 19.88) | .232 | 1.30 (−1.69 to 4.29) | .749 | 10.82 (0.87 to 20.76) | .139 |
| Tau PET | Parahippocampal cortex | −15.25 (−23.82 to −6.68) | **.008** | 1.89 (−1.04 to 4.81) | .560 | −13.36 (−21.20 to −5.52) | **.008** |
| Tau PET | Pars opercularis | 32.55 (10.17 to 54.93) | **.027** | 1.42 (−2.13 to 4.96) | .779 | 33.97 (11.77 to 56.17) | **.015** |
| Tau PET | Pars orbitalis | 12.59 (−4.74 to 29.92) | .403 | −1.75 (−4.63 to 1.13) | .573 | 10.84 (−6.31 to 27.98) | .474 |
| Tau PET | Pars triangularis | 16.24 (6.34 to 26.14) | **.012** | 0.62 (−1.61 to 2.86) | .831 | 16.87 (7.03 to 26.70) | **.008** |
| Tau PET | Pericalcarine cortex | 1.76 (−7.39 to 10.91) | .986 | 1.44 (−1.20 to 4.07) | .663 | 3.20 (−5.50 to 11.89) | .764 |
| Tau PET | Postcentral gyrus | 13.07 (5.14 to 21.00) | **.012** | −0.86 (−4.40 to 2.68) | .831 | 12.21 (5.17 to 19.26) | **.008** |
| Tau PET | Posterior cingulate | −0.61 (−34.03 to 32.81) | .986 | −0.08 (−3.28 to 3.12) | .961 | −0.69 (−34.00 to 32.61) | .999 |
| Tau PET | Precentral gyrus | 15.53 (3.84 to 27.22) | **.049** | 3.01 (−0.70 to 6.72) | .464 | 18.54 (7.32 to 29.75) | **.009** |
| Tau PET | Precuneus | 44.00 (−50.99 to 138.99) | .662 | 0.41 (−4.08 to 4.90) | .956 | 44.40 (−50.54 to 139.34) | .653 |
| Tau PET | Putamen | 1.86 (−11.86 to 15.58) | .986 | 0.70 (−1.89 to 3.29) | .831 | 2.56 (−10.90 to 16.02) | .991 |
| Tau PET | Rostral anterior cingulate | 10.68 (−2.21 to 23.57) | .312 | −0.16 (−2.35 to 2.03) | .956 | 10.52 (−2.20 to 23.24) | .293 |
| Tau PET | Rostral middle frontal | 16.38 (−0.22 to 32.98) | .186 | −1.84 (−4.74 to 1.06) | .560 | 14.54 (−1.51 to 30.60) | .249 |
| Tau PET | Superior frontal | 15.82 (−4.52 to 36.16) | .355 | −1.74 (−5.58 to 2.09) | .749 | 14.08 (−5.65 to 33.81) | .417 |
| Tau PET | Superior parietal | −11.28 (−29.40 to 6.84) | .490 | −1.09 (−5.06 to 2.89) | .831 | −12.37 (−30.02 to 5.28) | .417 |
| Tau PET | Superior temporal | 0.70 (−52.50 to 53.91) | .986 | 0.19 (−4.36 to 4.75) | .956 | 0.90 (−52.35 to 54.15) | .999 |
| Tau PET | Supramarginal gyrus | 20.09 (1.53 to 38.65) | .143 | −3.69 (−7.51 to 0.14) | .418 | 16.40 (−1.79 to 34.60) | .249 |
| Tau PET | Temporal pole | −6.23 (−28.94 to 16.48) | .917 | −1.96 (−4.45 to 0.53) | .464 | −8.19 (−30.63 to 14.26) | .764 |
| Tau PET | Thalamus | 5.47 (−3.33 to 14.26) | .490 | 1.64 (−0.75 to 4.02) | .560 | 7.10 (−1.39 to 15.60) | .293 |
| Tau PET | Transverse temporal | −1.32 (−11.94 to 9.29) | .986 | 2.54 (0.21 to 4.88) | .418 | 1.22 (−9.18 to 11.62) | .999 |
| Tau PET | Ventral diencephalon | −0.65 (−12.17 to 10.87) | .986 | 0.83 (−1.83 to 3.48) | .831 | 0.18 (−11.11 to 11.46) | .999 |
| Morphometry | Accumbens area | 3.61 (−1.27 to 8.50) | .805 | 2.59 (0.69 to 4.49) | .171 | 6.20 (1.60 to 10.80) | .177 |
| Morphometry | Amygdala | 1.87 (−4.23 to 7.97) | .990 | 2.38 (−1.00 to 5.76) | .780 | 4.25 (−1.89 to 10.38) | .614 |
| Morphometry | Banks of superior temporal sulcus | 0.92 (−4.42 to 6.26) | .990 | 0.29 (−1.62 to 2.20) | .985 | 1.21 (−3.89 to 6.31) | .902 |
| Morphometry | Caudal anterior cingulate | 5.77 (1.47 to 10.07) | .366 | 0.27 (−1.39 to 1.93) | .985 | 6.04 (2.05 to 10.02) | .132 |
| Morphometry | Caudal middle frontal | 1.15 (−3.99 to 6.29) | .990 | 0.24 (−1.47 to 1.95) | .985 | 1.38 (−3.53 to 6.29) | .902 |
| Morphometry | Caudate | 0.49 (−6.01 to 6.99) | .997 | 1.59 (−1.51 to 4.70) | .914 | 2.08 (−4.23 to 8.39) | .902 |
| Morphometry | Cuneus | −0.65 (−5.80 to 4.49) | .997 | 0.64 (−1.54 to 2.81) | .985 | −0.02 (−4.70 to 4.66) | .994 |
| Morphometry | Entorhinal cortex | 6.25 (0.69 to 11.81) | .389 | −0.61 (−2.53 to 1.31) | .985 | 5.64 (0.52 to 10.77) | .435 |
| Morphometry | Frontal pole | 0.22 (−4.22 to 4.67) | .997 | 1.11 (−1.44 to 3.67) | .967 | 1.34 (−2.45 to 5.13) | .902 |
| Morphometry | Fusiform gyrus | −1.61 (−7.24 to 4.02) | .990 | 2.65 (0.69 to 4.60) | .171 | 1.04 (−4.37 to 6.45) | .902 |
| Morphometry | Hippocampus | 4.69 (−0.12 to 9.49) | .587 | −0.60 (−2.84 to 1.63) | .985 | 4.08 (−0.25 to 8.41) | .509 |
| Morphometry | Inferior parietal | −0.20 (−5.48 to 5.07) | .997 | 1.27 (−0.86 to 3.41) | .840 | 1.07 (−3.90 to 6.04) | .902 |
| Morphometry | Inferior temporal | −3.89 (−8.75 to 0.97) | .805 | 2.89 (0.19 to 5.59) | .381 | −1.01 (−5.19 to 3.18) | .902 |
| Morphometry | Insula | 2.67 (−2.19 to 7.53) | .990 | −0.25 (−2.19 to 1.70) | .985 | 2.43 (−2.16 to 7.01) | .861 |
| Morphometry | Isthmus cingulate | −5.94 (−10.94 to −0.94) | .389 | 1.78 (0.20 to 3.36) | .380 | −4.16 (−8.96 to 0.65) | .509 |
| Morphometry | Lateral occipital | 2.14 (−2.67 to 6.95) | .990 | 1.02 (−0.60 to 2.64) | .828 | 3.16 (−1.42 to 7.73) | .614 |
| Morphometry | Lateral orbitofrontal | 2.07 (−2.18 to 6.32) | .990 | −0.26 (−2.25 to 1.74) | .985 | 1.81 (−2.10 to 5.72) | .902 |
| Morphometry | Lingual gyrus | −0.01 (−5.83 to 5.80) | .997 | 0.48 (−1.28 to 2.23) | .985 | 0.46 (−5.07 to 6.00) | .912 |
| Morphometry | Medial orbitofrontal | 0.77 (−3.76 to 5.30) | .990 | −0.27 (−2.37 to 1.83) | .985 | 0.51 (−3.58 to 4.59) | .902 |
| Morphometry | Middle temporal | 0.23 (−4.48 to 4.94) | .997 | 2.01 (−0.12 to 4.14) | .542 | 2.24 (−2.08 to 6.56) | .861 |
| Morphometry | Pallidum | −1.84 (−6.53 to 2.85) | .990 | 0.17 (−2.49 to 2.82) | .988 | −1.67 (−6.43 to 3.09) | .902 |
| Morphometry | Paracentral lobule | −1.69 (−6.84 to 3.45) | .990 | 1.21 (−0.65 to 3.07) | .828 | −0.48 (−5.41 to 4.44) | .912 |
| Morphometry | Parahippocampal cortex | 4.44 (−1.44 to 10.32) | .805 | 0.16 (−2.11 to 2.42) | .988 | 4.60 (−0.79 to 9.98) | .509 |
| Morphometry | Pars opercularis | −1.38 (−6.61 to 3.84) | .990 | 0.06 (−2.08 to 2.20) | .988 | −1.32 (−6.14 to 3.50) | .902 |
| Morphometry | Pars orbitalis | 1.13 (−3.58 to 5.83) | .990 | −1.82 (−4.39 to 0.74) | .780 | −0.69 (−4.63 to 3.25) | .902 |
| Morphometry | Pars triangularis | 0.09 (−4.90 to 5.07) | .997 | −0.03 (−2.30 to 2.24) | .988 | 0.05 (−4.51 to 4.62) | .994 |

| Modality | Region | Cohort difference, B (95% CI) | FDR p | PREVENT-AD, B (95% CI) | FDR p | UCLouvain, B (95% CI) | FDR p |
|---|---|---|---|---|---|---|---|
| Morphometry | Pericalcarine cortex | −1.65 (−7.56 to 4.27) | .990 | −0.03 (−1.94 to 1.89) | .988 | −1.68 (−7.25 to 3.90) | .902 |
| Morphometry | Postcentral gyrus | −1.07 (−6.94 to 4.80) | .990 | 0.01 (−1.77 to 1.79) | .988 | −1.05 (−6.83 to 4.72) | .902 |
| Morphometry | Posterior cingulate | −0.63 (−4.56 to 3.31) | .990 | 0.07 (−2.09 to 2.23) | .988 | −0.56 (−3.99 to 2.88) | .902 |
| Morphometry | Precentral gyrus | −1.34 (−7.08 to 4.39) | .990 | 0.26 (−1.44 to 1.95) | .985 | −1.09 (−6.67 to 4.50) | .902 |
| Morphometry | Precuneus | 0.05 (−4.61 to 4.70) | .997 | 0.64 (−1.26 to 2.53) | .985 | 0.68 (−3.70 to 5.07) | .902 |
| Morphometry | Putamen | 2.36 (−3.20 to 7.93) | .990 | 2.56 (−0.75 to 5.86) | .780 | 4.92 (−0.25 to 10.10) | .509 |
| Morphometry | Rostral anterior cingulate | 4.25 (−1.60 to 10.10) | .805 | 0.42 (−1.60 to 2.43) | .985 | 4.67 (−0.85 to 10.18) | .509 |
| Morphometry | Rostral middle frontal | −1.55 (−6.85 to 3.76) | .990 | 0.98 (−1.16 to 3.12) | .967 | −0.57 (−5.40 to 4.26) | .902 |
| Morphometry | Superior frontal | 0.46 (−4.66 to 5.58) | .997 | 0.21 (−1.63 to 2.06) | .985 | 0.67 (−4.21 to 5.54) | .902 |
| Morphometry | Superior parietal | 0.65 (−4.60 to 5.91) | .997 | 0.33 (−1.56 to 2.23) | .985 | 0.98 (−4.06 to 6.03) | .902 |
| Morphometry | Superior temporal | −2.26 (−7.01 to 2.49) | .990 | 0.69 (−1.44 to 2.82) | .985 | −1.57 (−6.22 to 3.08) | .902 |
| Morphometry | Supramarginal gyrus | 0.90 (−4.23 to 6.03) | .990 | 0.76 (−1.12 to 2.63) | .985 | 1.66 (−3.35 to 6.66) | .902 |
| Morphometry | Temporal pole | 1.89 (−2.96 to 6.74) | .990 | 1.39 (−1.03 to 3.80) | .840 | 3.28 (−0.98 to 7.53) | .611 |
| Morphometry | Thalamus | 3.37 (−3.34 to 10.08) | .990 | 0.61 (−1.73 to 2.95) | .985 | 3.98 (−2.87 to 10.83) | .820 |
| Morphometry | Transverse temporal | −1.64 (−7.02 to 3.74) | .990 | 0.86 (−0.86 to 2.58) | .914 | −0.78 (−5.89 to 4.33) | .902 |
| Morphometry | Ventral diencephalon | 1.72 (−3.56 to 6.99) | .990 | 1.98 (−0.79 to 4.76) | .780 | 3.70 (−1.54 to 8.94) | .614 |

**Note.** Sensitivity analysis excluding regional imaging values beyond ±3 within-cohort SD; tau-PET models also excluded extreme global cortical tau values, and subcortical-volume models excluded extreme eTIV values. B is the adjusted percentage-point change in odor identification score per 1 within-cohort SD higher biomarker value. Tau-PET models included global cortical tau and its interactions with cohort and amyloid status; subcortical-volume models included eTIV. FDR p values are corrected across all 42 regions separately by modality, group, and hypothesis. Cohort difference is UCLouvain minus PREVENT-AD.

#### Supplementary Table 10. Sensitivity analysis: atlas-wide amyloid-specific associations and biomarker-by-amyloid interactions

| Modality | Region | CN A−, B (95% CI) | FDR p | CN A+, B (95% CI) | FDR p | Amyloid interaction, B (95% CI) | FDR p |
|---|---|---|---|---|---|---|---|
| Tau PET | Accumbens area | 2.60 (0.55 to 4.66) | .185 | −0.01 (−4.95 to 4.92) | .996 | −2.61 (−7.71 to 2.48) | .711 |
| Tau PET | Amygdala | 3.25 (0.28 to 6.22) | .224 | −0.91 (−5.60 to 3.77) | .895 | −4.16 (−9.64 to 1.31) | .688 |
| Tau PET | Banks of superior temporal sulcus | 1.34 (−2.70 to 5.38) | .745 | −1.10 (−7.05 to 4.86) | .895 | −2.44 (−9.33 to 4.46) | .765 |
| Tau PET | Caudal anterior cingulate | −0.33 (−3.71 to 3.05) | .914 | 1.04 (−4.54 to 6.63) | .895 | 1.37 (−5.19 to 7.93) | .841 |
| Tau PET | Caudal middle frontal | 4.05 (−1.38 to 9.49) | .401 | 0.94 (−6.91 to 8.79) | .895 | −3.11 (−12.86 to 6.64) | .795 |
| Tau PET | Caudate | 2.58 (0.73 to 4.44) | .139 | −0.55 (−4.67 to 3.58) | .895 | −3.13 (−7.79 to 1.53) | .711 |
| Tau PET | Cuneus | 2.81 (−1.04 to 6.66) | .401 | 2.55 (−4.06 to 9.16) | .895 | −0.26 (−7.84 to 7.32) | .975 |
| Tau PET | Entorhinal cortex | 1.86 (−1.10 to 4.82) | .481 | −0.54 (−4.74 to 3.67) | .895 | −2.40 (−7.31 to 2.52) | .711 |
| Tau PET | Frontal pole | −1.07 (−4.24 to 2.10) | .745 | 1.30 (−4.82 to 7.42) | .895 | 2.37 (−4.41 to 9.14) | .765 |
| Tau PET | Fusiform gyrus | −0.71 (−5.00 to 3.57) | .891 | −3.79 (−8.82 to 1.23) | .528 | −3.08 (−9.63 to 3.47) | .711 |
| Tau PET | Hippocampus | 2.46 (0.30 to 4.63) | .219 | 1.41 (−1.93 to 4.76) | .895 | −1.05 (−5.03 to 2.94) | .801 |
| Tau PET | Inferior parietal | −2.85 (−6.86 to 1.16) | .403 | −8.73 (−18.78 to 1.32) | .488 | −5.88 (−16.66 to 4.90) | .711 |
| Tau PET | Inferior temporal | −0.08 (−5.20 to 5.05) | .977 | −7.42 (−14.98 to 0.13) | .488 | −7.35 (−16.38 to 1.68) | .661 |
| Tau PET | Insula | 4.04 (1.62 to 6.46) | **.049** | 0.70 (−6.70 to 8.11) | .895 | −3.34 (−11.24 to 4.57) | .711 |
| Tau PET | Isthmus cingulate | 1.30 (−2.10 to 4.70) | .745 | 0.48 (−3.30 to 4.25) | .895 | −0.82 (−6.04 to 4.39) | .894 |
| Tau PET | Lateral occipital | −0.81 (−4.72 to 3.11) | .872 | 3.76 (0.58 to 6.95) | .488 | 4.57 (−0.49 to 9.63) | .650 |
| Tau PET | Lateral orbitofrontal | 1.47 (−1.16 to 4.10) | .572 | −2.83 (−10.45 to 4.79) | .895 | −4.30 (−12.48 to 3.88) | .711 |
| Tau PET | Lingual gyrus | 1.09 (−1.85 to 4.04) | .745 | 3.46 (−0.58 to 7.51) | .488 | 2.37 (−2.73 to 7.48) | .711 |
| Tau PET | Medial orbitofrontal | 1.24 (−2.20 to 4.68) | .745 | 1.03 (−4.67 to 6.72) | .895 | −0.21 (−7.06 to 6.64) | .975 |
| Tau PET | Middle temporal | 4.86 (−1.65 to 11.37) | .401 | −5.53 (−14.83 to 3.76) | .724 | −10.39 (−21.74 to 0.96) | .650 |
| Tau PET | Pallidum | 2.58 (0.12 to 5.04) | .239 | −5.39 (−10.42 to −0.37) | .488 | −7.97 (−13.43 to −2.51) | .184 |
| Tau PET | Paracentral lobule | 1.26 (−1.51 to 4.03) | .709 | 2.99 (−2.66 to 8.65) | .776 | 1.73 (−4.45 to 7.92) | .801 |
| Tau PET | Parahippocampal cortex | 2.59 (−0.86 to 6.05) | .401 | −0.63 (−5.83 to 4.57) | .895 | −3.22 (−9.26 to 2.82) | .711 |
| Tau PET | Pars opercularis | 0.48 (−2.85 to 3.81) | .907 | 5.16 (−2.61 to 12.94) | .620 | 4.69 (−3.50 to 12.87) | .711 |
| Tau PET | Pars orbitalis | −0.98 (−4.10 to 2.14) | .751 | −1.02 (−6.85 to 4.81) | .895 | −0.04 (−6.48 to 6.41) | .991 |
| Tau PET | Pars triangularis | 0.48 (−2.31 to 3.26) | .891 | 4.82 (−0.45 to 10.09) | .488 | 4.35 (−1.54 to 10.23) | .688 |
| Tau PET | Pericalcarine cortex | 0.31 (−2.67 to 3.28) | .914 | 4.06 (−0.60 to 8.72) | .488 | 3.76 (−1.79 to 9.30) | .711 |
| Tau PET | Postcentral gyrus | −0.90 (−4.57 to 2.77) | .832 | 4.27 (−3.16 to 11.69) | .724 | 5.17 (−3.07 to 13.40) | .711 |
| Tau PET | Posterior cingulate | 0.30 (−3.62 to 4.22) | .924 | −0.57 (−5.84 to 4.70) | .895 | −0.87 (−7.38 to 5.64) | .900 |
| Tau PET | Precentral gyrus | 3.31 (−0.11 to 6.73) | .282 | 5.69 (−1.72 to 13.11) | .528 | 2.39 (−5.89 to 10.66) | .801 |
| Tau PET | Precuneus | 0.15 (−6.23 to 6.53) | .977 | 1.58 (−4.99 to 8.15) | .895 | 1.43 (−8.02 to 10.88) | .894 |
| Tau PET | Putamen | 2.28 (−0.16 to 4.71) | .282 | −2.51 (−7.82 to 2.79) | .821 | −4.79 (−10.59 to 1.02) | .661 |
| Tau PET | Rostral anterior cingulate | −0.57 (−2.92 to 1.78) | .832 | 2.57 (−2.45 to 7.60) | .776 | 3.14 (−2.41 to 8.69) | .711 |
| Tau PET | Rostral middle frontal | −1.94 (−4.74 to 0.86) | .403 | 2.01 (−6.34 to 10.35) | .895 | 3.95 (−4.89 to 12.78) | .711 |
| Tau PET | Superior frontal | −2.01 (−6.29 to 2.26) | .709 | 0.31 (−7.43 to 8.05) | .960 | 2.33 (−6.65 to 11.31) | .801 |
| Tau PET | Superior parietal | −1.24 (−4.35 to 1.87) | .745 | −2.16 (−12.04 to 7.71) | .895 | −0.92 (−11.21 to 9.37) | .936 |
| Tau PET | Superior temporal | 1.44 (−2.27 to 5.15) | .745 | −1.69 (−10.78 to 7.40) | .895 | −3.13 (−12.04 to 5.78) | .765 |
| Tau PET | Supramarginal gyrus | −3.46 (−7.46 to 0.54) | .343 | −1.31 (−10.23 to 7.61) | .895 | 2.15 (−7.44 to 11.73) | .839 |
| Tau PET | Temporal pole | −0.35 (−3.06 to 2.37) | .911 | −5.66 (−10.93 to −0.40) | .488 | −5.32 (−11.22 to 0.59) | .650 |
| Tau PET | Thalamus | 2.26 (0.29 to 4.23) | .219 | 1.74 (−4.23 to 7.72) | .895 | −0.51 (−6.63 to 5.60) | .936 |
| Tau PET | Transverse temporal | 1.66 (−0.62 to 3.95) | .401 | 3.84 (−0.96 to 8.65) | .528 | 2.18 (−2.87 to 7.24) | .711 |
| Tau PET | Ventral diencephalon | 2.69 (−0.15 to 5.52) | .282 | −3.30 (−7.82 to 1.22) | .530 | −5.99 (−11.24 to −0.73) | .542 |
| Morphometry | Accumbens area | 2.29 (0.65 to 3.92) | .265 | 5.68 (0.96 to 10.40) | .259 | 3.39 (−1.46 to 8.25) | .896 |
| Morphometry | Amygdala | 0.44 (−2.08 to 2.96) | .937 | 5.64 (0.54 to 10.73) | .281 | 5.20 (0.24 to 10.16) | .784 |
| Morphometry | Banks of superior temporal sulcus | 0.21 (−1.85 to 2.27) | .937 | 0.94 (−2.70 to 4.58) | .820 | 0.73 (−3.40 to 4.87) | .953 |
| Morphometry | Caudal anterior cingulate | 0.49 (−1.04 to 2.03) | .937 | 2.58 (−0.96 to 6.12) | .641 | 2.08 (−1.73 to 5.90) | .896 |
| Morphometry | Caudal middle frontal | 0.52 (−1.39 to 2.43) | .937 | 0.19 (−3.19 to 3.58) | .967 | −0.33 (−4.22 to 3.56) | .953 |
| Morphometry | Caudate | 0.96 (−1.82 to 3.73) | .937 | 3.24 (−3.07 to 9.55) | .758 | 2.28 (−4.13 to 8.70) | .923 |
| Morphometry | Cuneus | 0.21 (−1.92 to 2.34) | .937 | 1.29 (−2.90 to 5.47) | .820 | 1.08 (−3.59 to 5.75) | .953 |
| Morphometry | Entorhinal cortex | −0.38 (−2.26 to 1.51) | .937 | 2.60 (−1.70 to 6.89) | .739 | 2.98 (−1.82 to 7.77) | .896 |
| Morphometry | Frontal pole | 0.81 (−1.78 to 3.40) | .937 | 2.02 (−2.33 to 6.37) | .758 | 1.21 (−3.84 to 6.26) | .953 |
| Morphometry | Fusiform gyrus | 0.99 (−0.80 to 2.78) | .937 | 4.50 (0.77 to 8.23) | .259 | 3.51 (−0.51 to 7.53) | .784 |
| Morphometry | Hippocampus | −0.67 (−3.10 to 1.76) | .937 | 1.91 (−1.89 to 5.70) | .758 | 2.57 (−1.91 to 7.06) | .896 |
| Morphometry | Inferior parietal | 0.25 (−1.90 to 2.40) | .937 | 3.20 (−0.29 to 6.70) | .434 | 2.96 (−0.99 to 6.90) | .896 |

| Modality | Region | CN A−, B (95% CI) | FDR p | CN A+, B (95% CI) | FDR p | Amyloid interaction, B (95% CI) | FDR p |
|---|---|---|---|---|---|---|---|
| Morphometry | Inferior temporal | 2.21 (−0.64 to 5.05) | .937 | 1.87 (−2.02 to 5.77) | .758 | −0.33 (−4.97 to 4.31) | .953 |
| Morphometry | Insula | 0.00 (−2.08 to 2.09) | .997 | 0.60 (−2.52 to 3.71) | .867 | 0.59 (−2.90 to 4.09) | .953 |
| Morphometry | Isthmus cingulate | 0.85 (−0.83 to 2.53) | .937 | 0.99 (−2.57 to 4.55) | .820 | 0.14 (−3.75 to 4.03) | .953 |
| Morphometry | Lateral occipital | 0.17 (−0.98 to 1.32) | .937 | 4.15 (−0.02 to 8.32) | .358 | 3.98 (−0.30 to 8.26) | .784 |
| Morphometry | Lateral orbitofrontal | −0.53 (−2.40 to 1.34) | .937 | 1.65 (−2.48 to 5.78) | .820 | 2.19 (−2.17 to 6.54) | .906 |
| Morphometry | Lingual gyrus | −0.37 (−1.85 to 1.11) | .937 | 2.04 (−2.01 to 6.09) | .758 | 2.41 (−1.92 to 6.74) | .896 |
| Morphometry | Medial orbitofrontal | −0.37 (−2.44 to 1.71) | .937 | 0.34 (−3.23 to 3.91) | .942 | 0.70 (−3.30 to 4.71) | .953 |
| Morphometry | Middle temporal | 0.86 (−0.83 to 2.55) | .937 | 4.72 (0.37 to 9.07) | .281 | 3.86 (−0.65 to 8.38) | .784 |
| Morphometry | Pallidum | −0.39 (−3.05 to 2.27) | .937 | 0.71 (−3.57 to 5.00) | .867 | 1.10 (−3.05 to 5.26) | .953 |
| Morphometry | Paracentral lobule | 0.52 (−1.19 to 2.24) | .937 | 2.04 (−2.19 to 6.27) | .758 | 1.52 (−2.94 to 5.97) | .923 |
| Morphometry | Parahippocampal cortex | −1.32 (−3.03 to 0.38) | .937 | 6.44 (1.45 to 11.44) | .259 | 7.77 (2.37 to 13.16) | .207 |
| Morphometry | Pars opercularis | 0.43 (−1.53 to 2.38) | .937 | −1.41 (−5.82 to 3.00) | .820 | −1.84 (−6.63 to 2.96) | .923 |
| Morphometry | Pars orbitalis | −0.99 (−3.07 to 1.09) | .937 | −3.65 (−9.82 to 2.53) | .739 | −2.65 (−9.09 to 3.79) | .923 |
| Morphometry | Pars triangularis | 0.31 (−1.92 to 2.55) | .937 | −1.24 (−5.94 to 3.45) | .820 | −1.55 (−6.66 to 3.55) | .923 |
| Morphometry | Pericalcarine cortex | −0.26 (−2.40 to 1.87) | .937 | −0.08 (−3.51 to 3.35) | .967 | 0.18 (−3.90 to 4.26) | .953 |
| Morphometry | Postcentral gyrus | −0.32 (−1.99 to 1.35) | .937 | 0.20 (−4.11 to 4.52) | .967 | 0.52 (−4.03 to 5.07) | .953 |
| Morphometry | Posterior cingulate | 0.27 (−1.93 to 2.46) | .937 | −1.18 (−4.79 to 2.43) | .820 | −1.44 (−5.55 to 2.66) | .923 |
| Morphometry | Precentral gyrus | 0.10 (−1.65 to 1.85) | .966 | −0.08 (−3.81 to 3.65) | .967 | −0.18 (−4.23 to 3.87) | .953 |
| Morphometry | Precuneus | 0.46 (−1.21 to 2.13) | .937 | 1.09 (−3.31 to 5.49) | .820 | 0.63 (−3.99 to 5.25) | .953 |
| Morphometry | Putamen | 2.37 (−0.55 to 5.28) | .937 | 5.03 (−1.17 to 11.24) | .519 | 2.67 (−3.38 to 8.71) | .923 |
| Morphometry | Rostral anterior cingulate | 0.86 (−1.68 to 3.40) | .937 | 1.07 (−1.81 to 3.95) | .820 | 0.21 (−3.70 to 4.12) | .953 |
| Morphometry | Rostral middle frontal | 1.29 (−1.00 to 3.58) | .937 | −1.00 (−4.64 to 2.65) | .820 | −2.29 (−6.56 to 1.99) | .896 |
| Morphometry | Superior frontal | 0.62 (−1.45 to 2.70) | .937 | −0.51 (−3.54 to 2.51) | .867 | −1.13 (−4.69 to 2.43) | .923 |
| Morphometry | Superior parietal | −0.08 (−1.75 to 1.58) | .966 | 1.47 (−2.72 to 5.66) | .820 | 1.55 (−2.89 to 6.00) | .923 |
| Morphometry | Superior temporal | 0.06 (−1.73 to 1.86) | .966 | 0.91 (−3.84 to 5.65) | .867 | 0.84 (−3.83 to 5.51) | .953 |
| Morphometry | Supramarginal gyrus | 0.24 (−1.57 to 2.04) | .937 | 2.48 (−1.53 to 6.49) | .739 | 2.24 (−2.00 to 6.49) | .896 |
| Morphometry | Temporal pole | 1.15 (−1.16 to 3.46) | .937 | 2.95 (−1.93 to 7.83) | .739 | 1.80 (−3.59 to 7.19) | .923 |
| Morphometry | Thalamus | 0.85 (−1.54 to 3.25) | .937 | 1.50 (−3.38 to 6.37) | .820 | 0.64 (−4.36 to 5.64) | .953 |
| Morphometry | Transverse temporal | 0.63 (−0.90 to 2.16) | .937 | 0.49 (−3.72 to 4.71) | .928 | −0.13 (−4.64 to 4.37) | .953 |
| Morphometry | Ventral diencephalon | 1.67 (−0.79 to 4.12) | .937 | 5.38 (−0.88 to 11.64) | .482 | 3.71 (−2.03 to 9.45) | .896 |

**Note.** Sensitivity analysis excluding regional imaging values beyond ±3 within-cohort SD; tau-PET models also excluded extreme global cortical tau values, and subcortical-volume models excluded extreme eTIV values. B is the adjusted percentage-point change in odor identification score per 1 within-cohort SD higher biomarker value. Tau-PET models included global cortical tau and its interactions with cohort and amyloid status; subcortical-volume models included eTIV. FDR p values are corrected across all 42 regions separately by modality, group, and hypothesis. Amyloid interaction is CN A+ minus CN A−.

### Supplementary Table 11. Sensitivity analysis: atlas-wide biomarker-by-cohort-by-amyloid interactions

| Modality | Region | Cohort × amyloid interaction, B (95% CI) | p | FDR p | n |
|---|---|---|---|---|---|
| Tau PET | Accumbens area | −25.60 (−47.44 to −3.76) | **.022** | .229 | 241 |
| Tau PET | Amygdala | −24.66 (−66.71 to 17.39) | .249 | .531 | 237 |
| Tau PET | Banks of superior temporal sulcus | −48.93 (−90.43 to −7.43) | **.021** | .229 | 241 |
| Tau PET | Caudal anterior cingulate | −5.36 (−20.47 to 9.75) | .485 | .654 | 242 |
| Tau PET | Caudal middle frontal | 6.45 (−20.09 to 32.99) | .633 | .759 | 242 |
| Tau PET | Caudate | −14.30 (−30.66 to 2.07) | .087 | .454 | 240 |
| Tau PET | Cuneus | −7.25 (−23.09 to 8.59) | .368 | .638 | 239 |
| Tau PET | Entorhinal cortex | −17.00 (−34.40 to 0.40) | .055 | .388 | 240 |
| Tau PET | Frontal pole | 0.96 (−12.78 to 14.70) | .890 | .935 | 238 |
| Tau PET | Fusiform gyrus | −13.83 (−33.66 to 6.01) | .171 | .513 | 240 |
| Tau PET | Hippocampus | −17.30 (−38.00 to 3.40) | .101 | .472 | 238 |
| Tau PET | Inferior parietal | −1.71 (−48.02 to 44.60) | .942 | .964 | 242 |
| Tau PET | Inferior temporal | −19.73 (−49.93 to 10.46) | .199 | .531 | 241 |
| Tau PET | Insula | −23.09 (−46.48 to 0.30) | .053 | .388 | 241 |
| Tau PET | Isthmus cingulate | −35.80 (−65.45 to −6.14) | **.018** | .229 | 242 |
| Tau PET | Lateral occipital | 2.02 (−20.36 to 24.41) | .859 | .925 | 239 |
| Tau PET | Lateral orbitofrontal | −6.52 (−24.17 to 11.14) | .468 | .654 | 240 |
| Tau PET | Lingual gyrus | −9.02 (−21.89 to 3.85) | .168 | .513 | 238 |
| Tau PET | Medial orbitofrontal | −11.07 (−29.65 to 7.51) | .242 | .531 | 241 |
| Tau PET | Middle temporal | −23.34 (−62.13 to 15.45) | .237 | .531 | 241 |
| Tau PET | Pallidum | −8.66 (−29.00 to 11.69) | .403 | .638 | 239 |
| Tau PET | Paracentral lobule | −4.51 (−14.70 to 5.68) | .384 | .638 | 242 |
| Tau PET | Parahippocampal cortex | −6.67 (−29.72 to 16.37) | .569 | .724 | 240 |
| Tau PET | Pars opercularis | −10.30 (−36.29 to 15.70) | .436 | .638 | 241 |
| Tau PET | Pars orbitalis | 4.92 (−7.62 to 17.46) | .440 | .638 | 240 |
| Tau PET | Pars triangularis | 4.82 (−7.48 to 17.13) | .440 | .638 | 241 |
| Tau PET | Pericalcarine cortex | −5.52 (−16.96 to 5.93) | .343 | .638 | 239 |
| Tau PET | Postcentral gyrus | 2.56 (−8.72 to 13.84) | .656 | .765 | 241 |
| Tau PET | Posterior cingulate | −14.06 (−36.60 to 8.47) | .220 | .531 | 240 |
| Tau PET | Precentral gyrus | 0.34 (−14.34 to 15.02) | .964 | .964 | 241 |
| Tau PET | Precuneus | −63.46 (−112.83 to −14.10) | **.012** | .229 | 241 |
| Tau PET | Putamen | 5.70 (−10.87 to 22.26) | .499 | .654 | 241 |
| Tau PET | Rostral anterior cingulate | −7.69 (−21.69 to 6.31) | .280 | .560 | 241 |
| Tau PET | Rostral middle frontal | 3.42 (−9.77 to 16.62) | .610 | .753 | 241 |
| Tau PET | Superior frontal | −2.69 (−24.59 to 19.21) | .809 | .894 | 242 |
| Tau PET | Superior parietal | −24.96 (−58.89 to 8.97) | .149 | .513 | 240 |
| Tau PET | Superior temporal | −17.26 (−46.93 to 12.41) | .253 | .531 | 241 |
| Tau PET | Supramarginal gyrus | −18.23 (−44.22 to 7.76) | .168 | .513 | 242 |
| Tau PET | Temporal pole | −21.97 (−47.07 to 3.13) | .086 | .454 | 240 |
| Tau PET | Thalamus | −1.52 (−12.52 to 9.49) | .786 | .893 | 240 |
| Tau PET | Transverse temporal | −6.98 (−16.88 to 2.93) | .166 | .513 | 242 |
| Tau PET | Ventral diencephalon | −9.16 (−29.11 to 10.78) | .366 | .638 | 241 |
| Morphometry | Accumbens area | −0.37 (−13.24 to 12.51) | .955 | .996 | 246 |
| Morphometry | Amygdala | −0.98 (−14.13 to 12.17) | .884 | .996 | 247 |
| Morphometry | Banks of superior temporal sulcus | 2.82 (−7.33 to 12.98) | .585 | .994 | 248 |
| Morphometry | Caudal anterior cingulate | 4.80 (−3.41 to 13.01) | .251 | .752 | 248 |
| Morphometry | Caudal middle frontal | 9.05 (−1.56 to 19.66) | .094 | .743 | 247 |
| Morphometry | Caudate | 11.19 (−3.16 to 25.55) | .126 | .743 | 244 |
| Morphometry | Cuneus | −5.26 (−17.94 to 7.41) | .414 | .869 | 247 |
| Morphometry | Entorhinal cortex | 0.03 (−11.92 to 11.98) | .996 | .996 | 245 |
| Morphometry | Frontal pole | −1.21 (−13.69 to 11.28) | .849 | .996 | 246 |
| Morphometry | Fusiform gyrus | 0.47 (−10.29 to 11.23) | .932 | .996 | 246 |
| Morphometry | Hippocampus | −7.76 (−21.65 to 6.12) | .272 | .761 | 246 |
| Morphometry | Inferior parietal | 5.86 (−3.50 to 15.23) | .219 | .743 | 247 |

| Modality | Region | Cohort × amyloid interaction, B (95% CI) | p | FDR p | n |
|---|---|---|---|---|---|
| Morphometry | Inferior temporal | 1.68 (−9.26 to 12.62) | .762 | .996 | 246 |
| Morphometry | Insula | −1.05 (−14.46 to 12.37) | .878 | .996 | 247 |
| Morphometry | Isthmus cingulate | −7.29 (−17.11 to 2.53) | .145 | .743 | 248 |
| Morphometry | Lateral occipital | 7.91 (−1.87 to 17.68) | .112 | .743 | 247 |
| Morphometry | Lateral orbitofrontal | 6.43 (−7.61 to 20.47) | .368 | .869 | 247 |
| Morphometry | Lingual gyrus | 0.40 (−11.13 to 11.92) | .946 | .996 | 248 |
| Morphometry | Medial orbitofrontal | −0.29 (−10.50 to 9.93) | .956 | .996 | 246 |
| Morphometry | Middle temporal | 7.63 (−3.26 to 18.52) | .169 | .743 | 247 |
| Morphometry | Pallidum | −1.58 (−11.60 to 8.45) | .757 | .996 | 247 |
| Morphometry | Paracentral lobule | −0.17 (−14.02 to 13.69) | .981 | .996 | 248 |
| Morphometry | Parahippocampal cortex | 0.88 (−9.66 to 11.42) | .870 | .996 | 247 |
| Morphometry | Pars opercularis | 4.20 (−9.35 to 17.74) | .542 | .990 | 247 |
| Morphometry | Pars orbitalis | 4.88 (−9.10 to 18.87) | .492 | .962 | 246 |
| Morphometry | Pars triangularis | 17.12 (2.75 to 31.49) | **.020** | .431 | 247 |
| Morphometry | Pericalcarine cortex | −8.28 (−21.27 to 4.70) | .210 | .743 | 246 |
| Morphometry | Postcentral gyrus | 1.64 (−12.03 to 15.32) | .813 | .996 | 248 |
| Morphometry | Posterior cingulate | −17.45 (−33.38 to −1.51) | **.032** | .431 | 248 |
| Morphometry | Precentral gyrus | −0.14 (−13.45 to 13.17) | .984 | .996 | 248 |
| Morphometry | Precuneus | 1.97 (−8.46 to 12.39) | .711 | .996 | 246 |
| Morphometry | Putamen | 4.24 (−8.24 to 16.73) | .504 | .962 | 245 |
| Morphometry | Rostral anterior cingulate | 7.95 (−7.54 to 23.44) | .313 | .822 | 246 |
| Morphometry | Rostral middle frontal | 3.48 (−10.14 to 17.10) | .615 | .994 | 246 |
| Morphometry | Superior frontal | 5.99 (−7.67 to 19.65) | .389 | .869 | 248 |
| Morphometry | Superior parietal | 7.75 (−3.53 to 19.03) | .177 | .743 | 248 |
| Morphometry | Superior temporal | −2.98 (−17.09 to 11.13) | .678 | .996 | 247 |
| Morphometry | Supramarginal gyrus | 6.30 (−4.01 to 16.61) | .230 | .743 | 247 |
| Morphometry | Temporal pole | 7.42 (−10.00 to 24.84) | .402 | .869 | 244 |
| Morphometry | Thalamus | −13.88 (−26.66 to −1.09) | **.034** | .431 | 238 |
| Morphometry | Transverse temporal | −3.20 (−15.44 to 9.04) | .607 | .994 | 247 |
| Morphometry | Ventral diencephalon | −13.07 (−25.60 to −0.54) | **.041** | .431 | 248 |

Note. Sensitivity analysis excluding regional imaging values beyond ±3 within-cohort SD; tau-PET models also excluded extreme global cortical tau values, and subcortical-volume models excluded extreme eTIV values. B is the adjusted percentage-point change in odor identification score per 1 within-cohort SD higher biomarker value. Tau-PET models included global cortical tau and its interactions with cohort and amyloid status; subcortical-volume models included eTIV. FDR p values are corrected across all 42 regions separately by modality, group, and hypothesis. The reported coefficient is the biomarker-by-cohort-by-amyloid interaction. These models are exploratory.
